# Benefits and barriers to implementing precision preventive care: results of a national physician survey

**DOI:** 10.1101/2022.09.29.22280488

**Authors:** Jason L. Vassy, Benjamin J. Kerman, Elizabeth J. Harris, Amy A. Lemke, Marla L. Clayman, Ashley A. Antwi, Katharine MacIsaac, Thomas Yi, Charles A. Brunette

**Affiliations:** Harvard Medical School, Boston, Massachusetts, USA; Veterans Affairs Boston Healthcare System, Boston, Massachusetts, USA; Division of General Internal Medicine and Primary Care, Department of Medicine, Brigham and Women’s Hospital, Boston, Massachusetts, USA; Precision Population Health, Ariadne Labs, Boston, Massachusetts, USA; University of Louisville School of Medicine, Louisville, Kentucky, USA; UMass Chan Medical School, Department of Population and Quantitative Health Sciences, Worcester, Massachusetts, USA; Edith Nourse Rogers Memorial Veterans’ Hospital, Bedford, Massachusetts, USA

**Keywords:** Preventive medicine, polygenic risk scores, primary care, physician survey

## Abstract

**Background:** Clinical implementation of polygenic risk scores (PRS) for precision prevention depends on the utility and barriers primary care physicians (PCPs) perceive to their use.

**Methods:** An online survey asked PCPs in a national database about the clinical utility of PRS they perceived for categories of medical decision-making and perceived benefits of and barriers to that use. Latent class analysis (LCA) was used to identify subgroups of PCPs based on response patterns.

**Results:** Among 367 respondents (email open rate 10.8%; participation rate 96.3%; completion rate 93.1%), mean (SD) age was 54.9 (12.9) years, 137 (37.3%) were female, and mean (SD) time since medical school graduation was 27.2 (13.3) years. Respondents reported greater perceived utility for more clinical action (e.g., earlier or more intensive screening, preventive medications, or lifestyle modification) for patients with high-risk PRS than for delayed or discontinued prevention actions for low-risk patients (*p*<0.001). Respondents most often chose out-of-pocket costs (48%), lack of clinical guidelines (24%), and patient insurance discrimination concerns (22%) as extreme barriers to PRS implementation. LCA identified 3 subclasses of respondents. *Skeptics* (n=83, 22.6%) endorsed less agreement with individual clinical utilities, saw patient anxiety and insurance discrimination as significant barriers, and agreed less often that PRS could help patients make better health decisions. *Learners* (n=134, 36.5%) and *enthusiasts* (n=150, 40.9%) expressed similar levels of agreement that PRS had utility for preventive actions and that PRS could be useful for patient decision-making. Compared with *enthusiasts*, however, *learners* perceived greater barriers to the clinical use of PRS.

**Conclusion:** PCPs generally endorsed using PRS to guide medical decision-making about preventive care, with a preference for more clinical action over less. Barriers identified suggest interventions to address the needs and concerns of PCPs along the spectrum of acceptance and uptake.

## INTRODUCTION

Polygenic risk scores (PRS) have emerged as a promising tool for disease risk stratification in clinical medicine.^1–3^ Built on genome-wide association study data, a PRS aggregates small effect sizes from dozens to millions of variants from across the genome to convey a measure of an individual’s genetic predisposition to a given disease from common genetic variation. For some diseases, a high PRS value indicates a risk equivalent to that of high-penetrance, single-gene variants associated with monogenic disease.^4–6^ Advances in the diversity of available datasets and in computational methods are improving the accuracy of PRS in populations of diverse genetic ancestry.^7–9^

As a result of these developments, PRS are now being considered for implementation into clinical medicine. Clinical trials and implementation projects are underway,^10–14^ and several commercial laboratories already offer PRS products for clinical use.^15–17^ Most attention has focused on the potential for PRS to identify high-risk subgroups of patients for whom targeted interventions might improve health outcomes through earlier prevention, detection, or treatment;^4,10,18^ the potential for PRS to identify low-risk subgroups for whom certain preventive measures might be appropriately deferred remains more theoretical.^19,20^

Whereas preventive medicine is most often the domain of primary care, the successful implementation of PRS for risk stratification and prevention will depend on the clinical utility and barriers that primary care physicians (PCPs) perceive to their use. Given the increasing interest in the clinical application of PRS in preventive medicine contexts, it will be important to understand the perspectives of these frontline clinicians. To that end, we fielded a web-based national survey of U.S. PCPs about the potential clinical utility and barriers they perceive to the use of PRS in preventive care.

## METHODS

The study was approved by the Harvard Longwood Campus Institutional Review Board (Protocol #20-2098). We followed the Checklist for Reporting Results of Internet E-Surveys (CHERRIES) in presenting this research (Supplement).^21,22^

### Population and sampling strategy

We defined the target population as PCPs who care for adult patients, including physicians practicing family medicine, general practice, or internal medicine. We worked with database licensee IQVIA^®^ to recruit respondents from the *ONEKEY* national physician database of more than 250,000 active physicians who have opted in to receiving email survey invitations. The database includes demographic, training, and practice-related data from the American Medical Association (AMA) Physician Masterfile and other sources.^23^

### Recruitment

IQVIA sent email invitations with unique web links to the survey to a random sample of 27,000 eligible PCPs from the database. These emails described the questionnaire as an 8-10 minute survey about precision prevention in primary care using genetic risk scores. Recipients were able to opt out of further contact using a link in the invitation email. Respondents could access the survey through a unique web link, hosted by the Qualtrics^XM^ Survey Tool (Qualtrics, Provo, UT). The first email invitation was sent April 18, 2021, and offered respondents a $25 Amazon gift card for completing the survey. On April 27, 2021, a second email offering a $50 Amazon gift card was sent to PCPs who had opened the first email but not yet accessed the survey link from the first email. The survey was closed on August 27, 2021.

### Survey measures

The survey consisted of two major sections (Supplement). First, after a brief introduction to PRS, a set of clinical case scenarios assessed physicians’ medical decision-making with PRS results for atherosclerotic cardiovascular disease prevention and prostate cancer screening; we have previously reported the results of those survey items.^22^ The next section assessed PCPs’ perceived benefits and barriers to PRS testing, using questions adapted from prior PCP surveys on implementation of genetic and genomic medicine technologies. Specifically, PCP perceived utility of PRS was measured on a 5-point Likert scale (“strongly agree” to “strongly agree”) with questions asking PCPs whether they would use PRS to identify high-risk patients suitable for three “earlier or more intensive” actions: 1) disease screening procedures (e.g. mammography, colonoscopy), 2) recommendations for preventive medications (e.g. statins, aspirin, tamoxifen), and 3) recommendations for lifestyle modifications (e.g. weight loss and smoking cessation).^24–26^ Similarly, PCPs were asked whether they would use PRS to identify low-risk patients who might be able to “delay or discontinue” these three categories of preventive actions. Perceived benefits of PRS, including improvements in patient and provider decision making and patient health outcomes, and PCP confidence in their ability to use PRS were assessed with questions adapted from a prior survey.^27^ Perceived barriers to using PRS in clinical practice were assessed by asking PCPs to rate on a 4-point Likert scale from “not a barrier” to “extreme barrier” eight potential implementation barriers adapted from Mikat-Stevens and Lemke.^27,28^ The final questions asked respondents about any prior genetics education beyond the typical medical school curriculum^29^ and self-reported race and ethnicity using U.S. Census categories. Other respondent characteristics were obtained by linking individual survey responses to the IQVIA *ONEKEY* database: age, years in practice, gender, practice specialty, practice size, and geography by state. Data validation, assessment of data completion and careless responding, and the development of composite scores are described in the **Supplemental Methods**.

### Analysis

Analyses were conducted in R (v4.0.3, R Foundation, Vienna, Austria) and Stata (v17.0, StataCorp, College Station, TX, USA). Respondent characteristics and survey responses are presented descriptively. All structural modeling and other analyses are presented with relevant test statistics, *p*-values, and confidence intervals where appropriate. To identify subgroups of respondents based on similarities and differences in response patterns, we used latent class analysis (LCA), an exploratory latent variable modeling method that leverages data patterns to indirectly measure a categorical latent construct composed of distinct homogenous classes.^30^ The final indicator set included seven dichotomous items, and examination of model fit and interpretability favored a 3-class model. Additional details are included in the **Supplemental Methods**.

## RESULTS

### Participant characteristics

Of 25,803 physicians who received the email invitation without bounceback, 2,776 opened the email (email open rate 10.8%) and 409 participants clicked the hyperlink to view the survey (survey view rate 409/2,776, 14.7%). Of PCPs who viewed the survey, 394 consented to study participation (participation rate 394/409, 96.3%) and 367 completed all questions (completion rate 366/394, 93.1%). Among these 367 respondents, 232 (63.2%), 73 (19.9%), 12 (3.2%), and 5 (1.4%) self-reported white, Asian, Black/African-American and Native Hawaiian/Other Pacific Islander race, respectively; 15 (4.1%) self-reported Hispanic/Latinx ethnicity. The majority (329, 89.6%) reported no additional genetics training beyond medical school. Respondents had similar characteristics to non-respondents (**Supplemental Table 1**).

### Perceived utility of PRS for preventive actions

Figure 1. shows PCPs’ reported likelihood of using PRS to identify patients for whom they might change their medical decision-making about categories of preventive actions. The majority of PCPs (63%-93%) somewhat or strongly agreed that they would use PRS for all clinical actions queried except for using a low-risk PRS to identify patients who might be able to delay or discontinue recommendations for lifestyle modification (41% somewhat/strongly agreeing). Across composite scores associated with the 3 classes of preventive actions (disease screening procedures, preventive medications, and lifestyle modification), respondents endorsed stronger preference for taking earlier or more intensive action for high-risk patients than with delaying or discontinuing action for low-risk patients (all Wilcoxon signed rank test *p*<0.001 across composite and pairwise comparisons, **Supplemental Table 2**).

**Figure 1.**
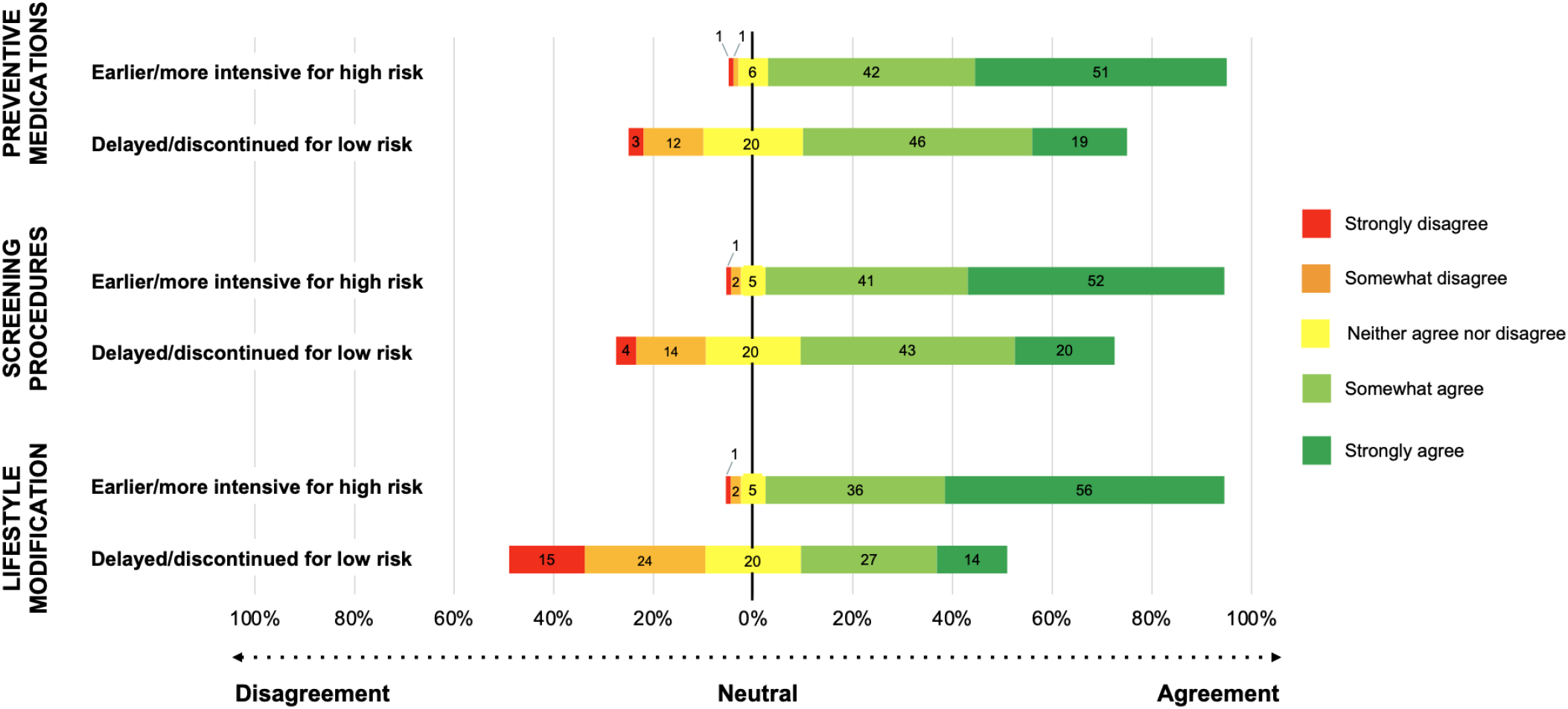
Perceived utility of PRS for preventive actions. Data indicate respondent agreement that they would use PRS in their clinical practice to identify high-risk patients who might need earlier/more intensive preventive action in three categories (preventive medications, screening procedures, and lifestyle modification) or to identify low-risk patients who might be able to delay/discontinue these preventive actions. Examples given were statins, aspirin, tamoxifen (preventive medications); mammography and colonoscopy (screening procedures); and weight loss and smoking cessation (lifestyle recommendations).

### Perceived benefits of PRS

The large majority of respondents agreed that PRS could help them (89% somewhat/strongly agreeing) and their patients (91% somewhat/strongly agreeing) improve their medical decision-making (**Figure 2**). A smaller majority (77%) somewhat or strongly agreed that PRS could help improve their patients’ health outcomes. **Supplemental Figure 1** shows respondent interest in using PRS for specific diseases, ranging from 88%-91% agreement for prostate, colorectal, and breast cancer to 48%-52% agreement for obesity, atrial fibrillation, and depression.

**Figure 2.**
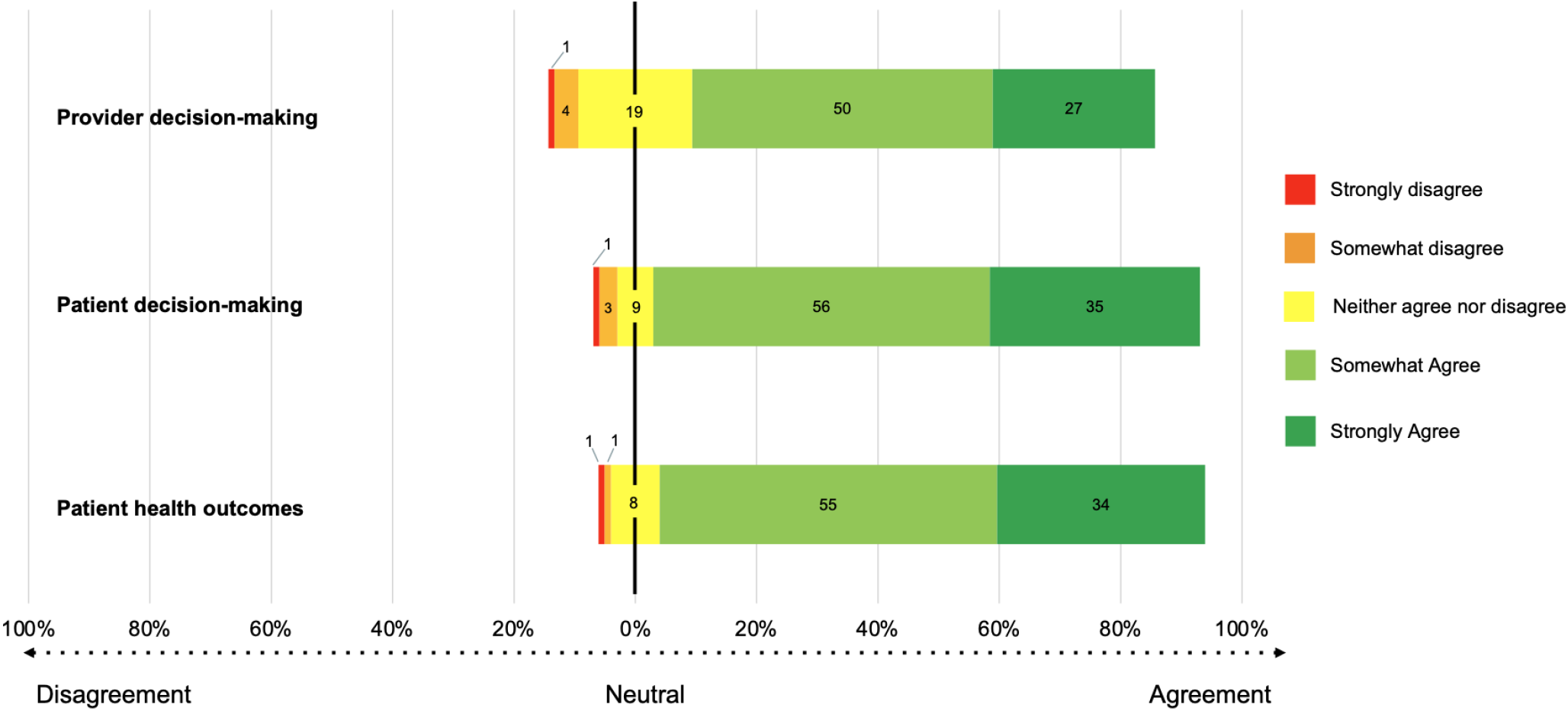
Perceived benefits of PRS. Data are respondent agreement with the statements *Genetic risk scores could help improve my medical decision-making in the care of patients*; *Genetic risk scores could help my patients make better decisions about their healthcare*; and *Genetic risk scores could improve my patients’ health outcomes*.

### Perceived barriers to PRS implementation

Figure 3. shows the significance respondents ascribed to potential barriers to using PRS in practice. Of the eight items, possible out-of-pocket cost to patients was most often chosen as a moderate or extreme barrier (86%), while insufficient time to explain PRS to patients was chosen least often (34%). All other potential barriers were chosen by the majority of respondents (51%-66%) as moderate or extreme barriers. Only 42% somewhat or strongly agreed with that statement “I am confident in my ability to use genetic risk score results.”

**Figure 3.**
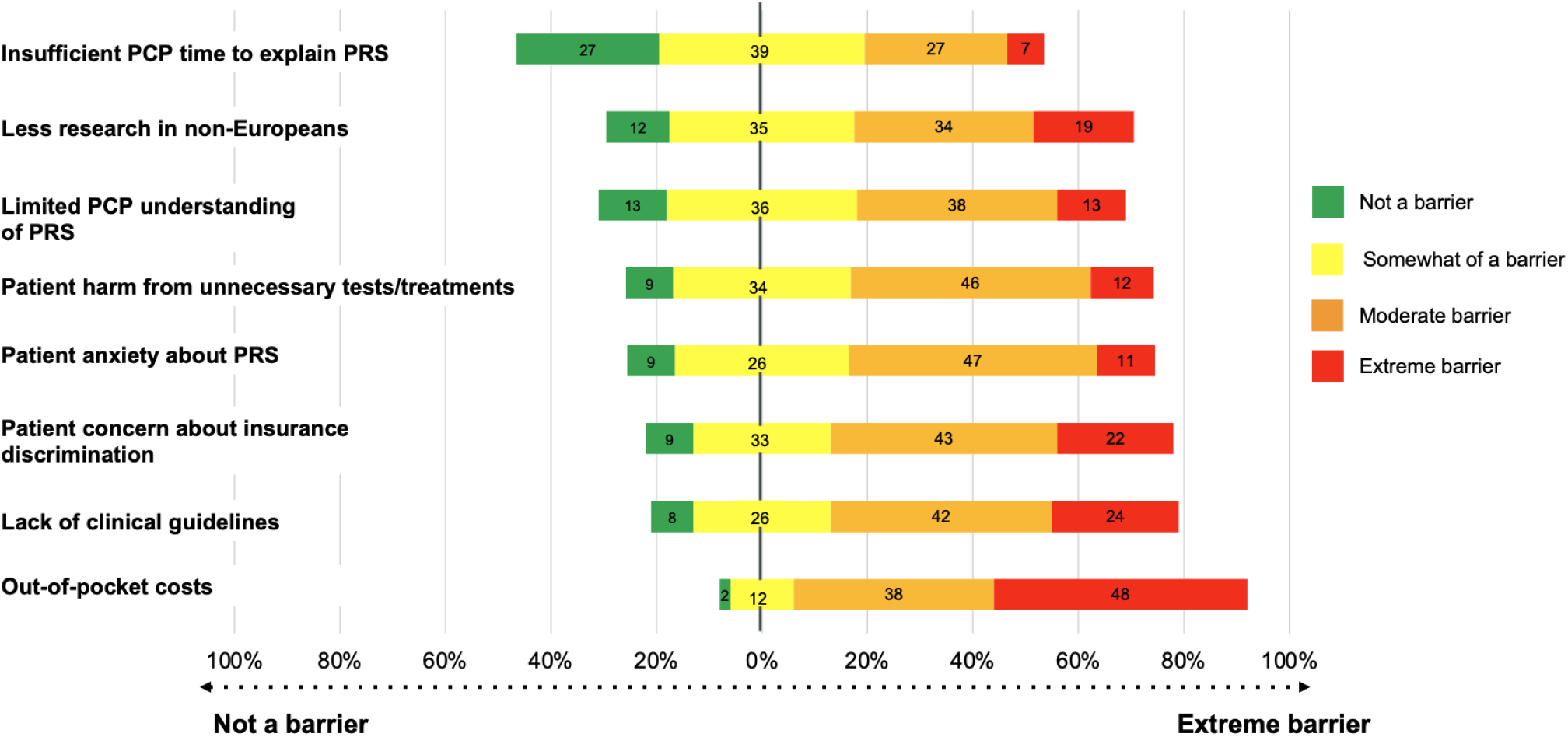
Perceived barriers to PRS implementation. Data are respondent ratings of the significance of each item as a barrier to using PRS in clinical practice.

### Latent class structure

A detailed description of LCA model fitting is described in the **Supplemental Results. Table 1** presents predicted class prevalences with 95% confidence intervals for the selected 3-class solution. Item-response probabilities associated with the final LCA model are presented in **Supplemental Table 3**.

*Class 1 (Skeptics):* Nearly one-quarter of survey response patterns (22.8%, 95% CI 16.9% to 30.0%) were characterized by high endorsement probabilities for barriers to PRS use, including that PRS would result in unnecessary testing or treatment (0.799, 95% CI 0.680, 0.882) and are less studied in non-European populations (0.790, 95% CI 0.666, 0.876). These patterns suggested only moderate agreement that PRS could improve clinical decision-making (0.585, 95% CI 0.463, 0.698) and endorsed uncertainty around or disagreement with the statements that PRS could improve patient health outcomes (0.148, 95% CI 0.041, 0.410) and that respondents could confidently use PRS in medical practice (0.084, 95% CI 0.036, 0.183). We labeled this latent class *skeptics*.

*Class 2 (Learners):* Approximately one-third of response patterns (33.5%, 95% CI 26.0% to 42.0%) were also characterized by high endorsement probabilities for barriers to PRS, but their response patterns were distinguished by very high endorsement of the statements that they had insufficient understanding about how to use PRS (0.987, 95% 0.889, 1.000) but that PRS could improve both medical decision-making (0.977, 95% CI 0.885, 0.996) and patient outcomes (0.999, 95% CI 0.999, 1.000). These patterns were also associated with uncertainty or disagreement that respondents could confidently use PRS in their clinical practice (0.329, 95% CI 0.241, 0.432). We labeled this latent class *learners*.

*Class 3 (Enthusiasts):* The remainder of response patterns (43.7%, 95% CI 36.2% to 51.6%) aligned with fewer endorsed barriers to using PRS (all item-response probabilities <0.350) and suggested high to very high agreement that PRS could improve medical decision-making (0.978, 95% CI 0.910, 0.995) and patient outcomes (0.908, 95% CI 0.830, 0.952). These respondents were confident in their ability to use PRS results in practice (0.670, 95% 0.577, 0.752). Given the inclination toward fewer barriers, heightened utility, and confident use of PRS, we labeled this latent class *enthusiasts*.

### Characteristics of latent classes

**Table 1** shows characteristics by latent class. High agreement was observed between estimated class prevalences and predicted class memberships among respondents: 83 (22.6%) *skeptics*, 134 (36.5%) *learners*, and 150 (40.9%) *enthusiasts*. Between-class differences in demographics, professional history, and practice location were not statistically significant. Slightly higher proportions of female respondents were observed among *skeptics* (38.6%) and *learners* (44.0%) compared to *enthusiasts* (30.7%, *p*=0.065). *Enthusiasts* were more likely to report additional genetics training beyond medical school (14.7%) than *learners* (6.7%) or *skeptics* (8.4%, *p*=0.073).

**TABLE 1.**
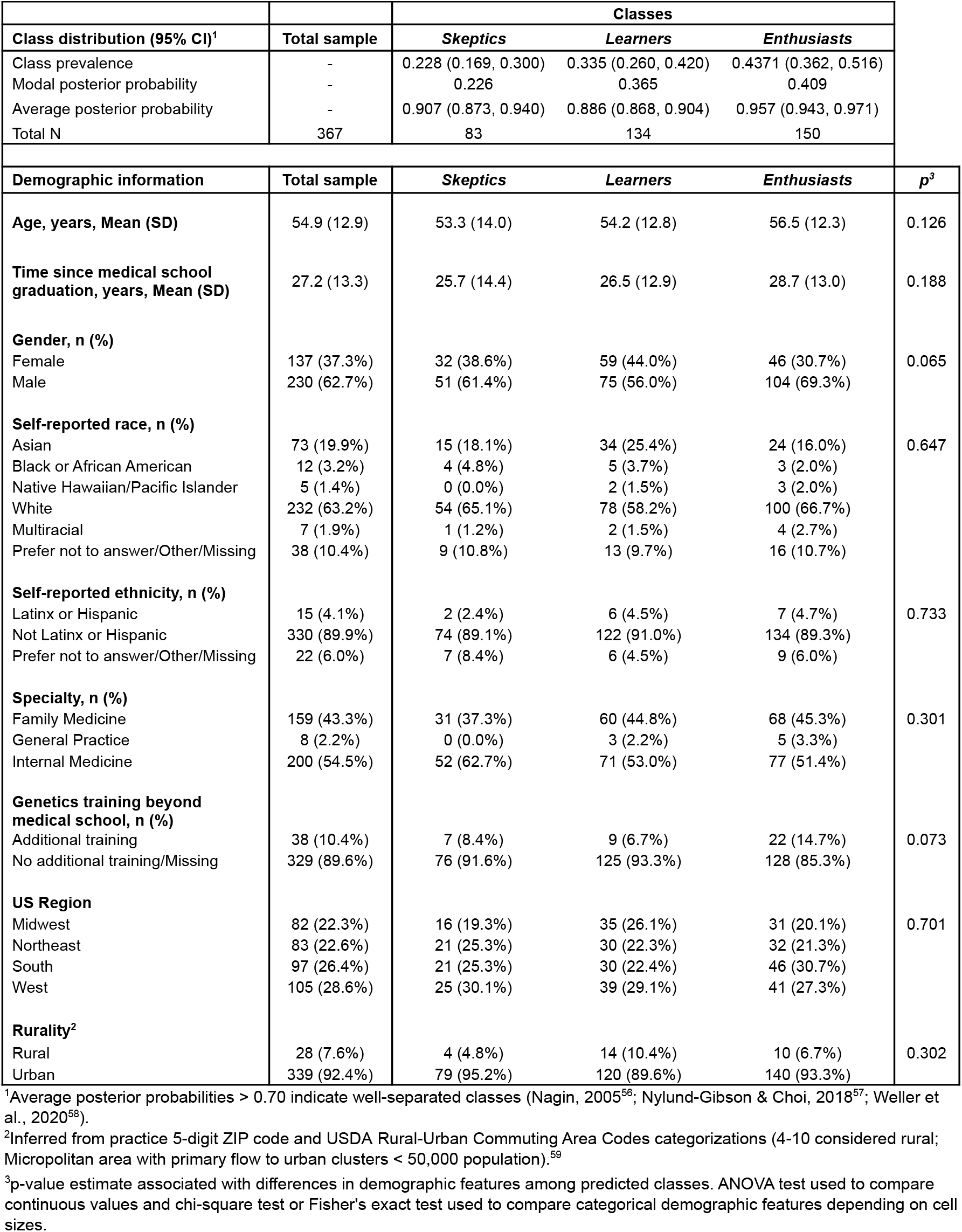
Latent class prevalence and participant characteristics.

Observed endorsements of items not included in LCA modeling aligned as expected with predicted survey response patterns and class memberships (**Figure 4**). S*keptics* consistently endorsed lower agreement with individual items regarding the use of PRS for earlier or delayed preventive action, expressed the most concern that patient anxiety and insurance discrimination were also significant barriers, were less likely to agree that PRS could help patients make better health care decisions. *Learners* and *enthusiasts* generally expressed higher levels of agreement that PRS could be used for either earlier or delayed clinical action and that PRS could help patient decision-making. Compared to *enthusiasts*, however, *learners* acknowledged greater barriers to using PRS across all items and were most likely to consider absence of guidelines as the primary barrier to using PRS in practice. All three classes noted out-of-pocket costs as a barrier.

**Figure 4.**
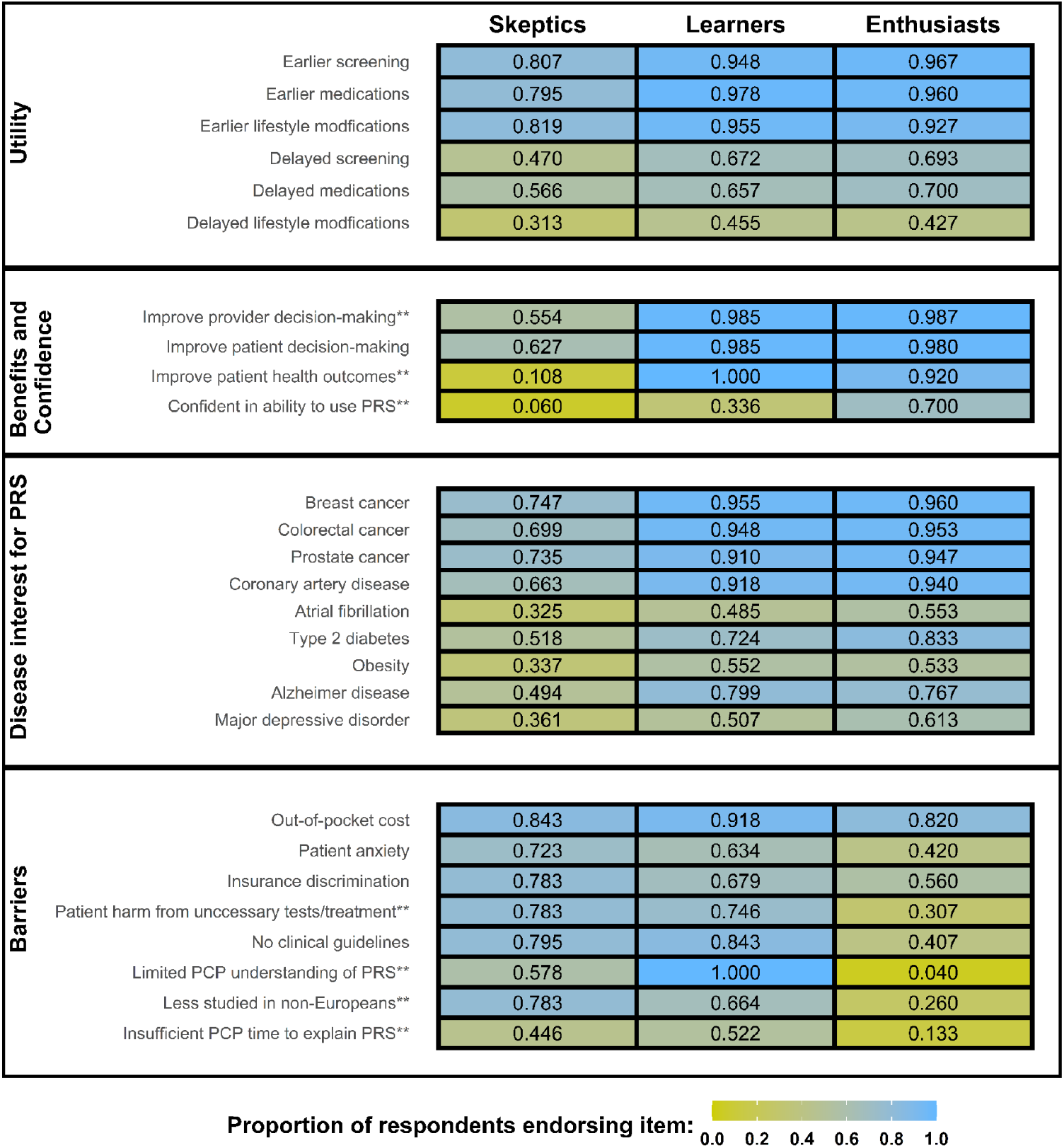
Observed survey item endorsements by predicted class membership. Data are proportions of respondents endorsing greater agreement or barrier significance for selected dichotomized items. **Item included in final latent class analysis model.

Between-class differences were also observed for both early preventive action composite scores for high-risk PRS (Kruskal-Wallis rank sum test *p*<0.001) and delayed action composite scores for low-risk PRS (*p*=0.005). Pairwise comparisons demonstrated less overall propensity to favor earlier or delayed preventive action among *skeptics* compared to *learners* (early action *p*<0.001; delayed action *p*=0.005) and *enthusiasts* (early action *p*<0.001; delayed action *p*=0.005). No statistically significant median differences were observed between *learners* and *enthusiasts* (early action *p*=0.081; delayed action *p*=0.848). Within all classes, respondents favored more clinical action for high-risk PRS results over less action for low-risk results (*skeptics p*<0.001; *learners p*<0.001; *enthusiasts p*<0.001).

## DISCUSSION

In the largest national physician survey about PRS to date, we observed general endorsement among PCPs for using PRS to guide medical decision-making about preventive medicine actions. Respondents favored earlier action for high-risk individuals more than delayed action for low-risk individuals. They also endorsed certain barriers to the clinical implementation of PRS, and fewer than half reported confidence in their ability to use PRS. Latent class analysis suggested 3 classes of PCPs with regard to the utility and barriers they perceived for PRS: *skeptics* who heavily weighted the barriers to and potential harms of PRS; *learners* who perceived both barriers and benefits to implementation; and *enthusiasts* who perceived benefit and self-confidence in the use of PRS. These observations suggest policy and system interventions to optimize the implementation of PRS into preventive care.

A primary focus of the survey was whether and how PCPs anticipated using PRS to make medical decisions. The majority reported they would use PRS results to change their recommendations for preventive actions, suggesting some degree of PCP acceptance of PRS as actionable results. However, among respondents overall and within each latent class, we observed a clear preference for doing more for high-risk results than doing less for low-risk results. This pattern might represent commission bias among respondents, an inclination towards clinical action even when inaction might result in equivalent or more favorable outcomes.^31,32^ It might also reflect their concerns, observed in studies of risk-stratified breast cancer screening,^33–35^ about whether PRS have sufficient accuracy to justify reducing recommended screenings at the risk of delaying detection. Current or proposed implementation of PRS also reflects this bias towards action. PRS for breast cancer and coronary artery disease are proposed as tools (or *risk-enhancing factors*) to identify high-risk patients eligible for screening and statin therapy, respectively, that traditional risk stratification methods miss.^15,18^ Whether PRS are used in the future as *protective factors* to help patients safely defer preventive care will require strong evidence of benefit and effective communication to PCPs and patients that doing so is to reduce preventive care-related risks and not about rationing health care.^34,36^

The barriers to clinical PRS implementation that PCPs selected are consistent with prior studies, in which common concerns about the use of PRS included lack of knowledge or confidence,^37–40^ insufficient evidence or guidelines to support their use,^35,38,39,41–43^ and insurance discrimination and other ethical issues.^26,37,39,43,44^ In the present survey, it is noteworthy that out-of-pocket costs and concerns about patient insurance discrimination were among the three most commonly selected barriers to PRS implementation. Respondents brought their own information and perceptions in these responses, as the survey did not inform them about the cost of PRS testing or about the Genetic Information Nondiscrimination Act of 2008, which prohibits discrimination on the basis of genetic information in employment and health insurance, but not in life or disability insurance.^45^ The majority (73%) of respondents categorized insufficient time to explain PRS to patients as at least somewhat of a barrier; however, given concerns about PCP burnout,^46,47^ it is striking that this was the least often selected potential barrier. Barrier response patterns varied among the 3 latent classes of PCP identified from the data. *Enthusiasts* generally endorsed few barriers to the use of PRS beyond out-of-pocket costs, while *skeptics* and *learners* also often identified patient anxiety, unintended harms, insufficient understanding, and the lack of studies among non-European populations as concerns. *Learners* were distinguished from *skeptics* in their agreement that PRS might help improve health decision-making and outcomes.

These findings point to a research and policy agenda to promote the uptake of PRS into preventive care. First, additional research is needed to clarify the benefits and harms of PRS-informed risk stratification. The necessary evidence will need to detail the specific diseases and patient populations for which PRS might have the most favorable benefit-to-harm ratio. This evidence will likely come from a combination of clinical epidemiology and modeling studies, select randomized trials, and clinical implementation projects and should prioritize important clinical outcomes. Second, this evidence should inform the development of clinical guidelines from relevant professional organizations for the appropriate use of PRS in clinical care. Given the proliferation of PRS, it will be beneficial for such organizations to state proactively the benchmarks that a PRS must meet to prove beneficial for the preventive care of their diseases of interest, even if they do not find the evidence supports such use at the present time. If guidelines do endorse the use of PRS, they should include both recommendations for appropriate management and identification of what they would consider inappropriate management. Such guidelines would address the learners in our study who wanted guidance on how to use PRS and the skeptics who were concerned about their harms and unnecessary tests and treatments that might result. They could also be incorporated into clinical decision support tools within the electronic medical record to promote appropriate usage. Third, professional organizations and healthcare systems should summarize information about cost and insurance discrimination to PCPs ordering PRS testing. This information is complex, dynamic, and variable by country and U.S. state,^48,49^ but providers and patients will need accurate information to make informed decisions.

One limitation of this study is its hypothetical nature. Few respondents likely have actual clinical experience with PRS, and their responses might not reflect their actual practice patterns as PRS become more commonplace. Second, only 10.8% of invited PCPs opened the survey invitation email. This rate compares favorably with those of other large national physician surveys,^50,51^ including those about genetic testing and precision medicine.^26,40,41,52–54^ Response rate is not the best indicator of non-response bias,^55^ and our comparison of the sample to the target population gives some support for its representativeness. The invitation described the study as a “survey on precision prevention in primary care.” Respondents may have had more interest in PRS than average PCPs, but we nonetheless observed a range of positive and negative attitudes.

In conclusion, data from the largest national survey about PRS to date suggest that PCPs see utility in using PRS in preventive medicine. The pressing needs for the future of PRS implementation include evidence and guidelines to support their appropriate use by PCPs and clarity around their financial consequences.

## Supporting information

Survey instrument

CHERRIES checklist

## Data Availability

All data produced in the present study are available upon reasonable request to the authors.

## Authors contributions

CRediT category: Conceptualization: JLV, BJK; Data curation: EJH, KI, TY; Formal analysis: CAB; Funding acquisition: JLV, BJK; Investigation: JLV, BJK, EJH; Methodology: JLV, BJK, EJH, AAL, MLC, JLV; Project administration: EJH, AAA; Software: EJH, TY, CAB; Supervision: JLV; Validation: EJH, CAB; Visualization: JLV, BJK, EJH, KI, TY, CAB; Writing - original draft: JLV, CAB; Writing - review & editing: JLV, BJK, EJH, AAL, MLC, AAA, KI, TY, CAB.

## SUPPLEMENTAL METHODS

### Data validation

Prior to analysis, survey data were assessed for response validity. Duplicate responses were identified via a unique survey link identifier. In the case of multiple responses associated with the same identifier, only the initial response was considered for analysis. To promote item response, each of the first ten questions (all except those on respondent characteristics) required a selection before the respondent could proceed. Respondents provided an email address to request the gift card incentive on the final page of the survey, only after responding to the primary survey items. After survey submission, we further assessed the dataset for invalid responses using common metrics for careless or invalid responding.^60^ We considered potentially invalid responses as those with: 1) total survey response times less than 4 minutes (bottom 5% of all survey times), 2) longstrings (selection of the same response choice over multiple survey items) in the top 90th percentile across all items, and 3) status as a multivariate outlier using Mahalanobis distance over all survey items and across at least three individual question sets. A total of 29 participants met at least one criterion for potentially careless or invalid responding.

Seventeen participants completed the survey in less than 4 minutes, 1 participant recorded longstrings in the top 90th percentile, 8 participants were considered multivariate outliers, and 3 participants met two or more criteria for careless responding. Sensitivity analyses removing these participants from the dataset were conducted. Conclusions using the remaining 338 respondents across all structural and analytic modeling were consistent with full sample assessment (sensitivity analysis output available upon request). Response validity was assessed in R (v4.0.3) with the *careless* package (v1.2.1).

### Structural modeling

To assess survey participants’ overall propensity to endorse clinical actions in the presence of high- or low-risk PRS, we developed composite scores of both early and delayed action items related to the stem “*In my clinical practice, I would use genetic risk scores to…*” Each composite included the sum of 3 items related to either earlier or delayed screening, more intensive or delayed medication recommendations, and more intensive or delayed recommendations for lifestyle modification. Composite scores were considered representative of the larger constructs of early/more clinical action and delayed/less clinical action. Scores ranged from 3 to 15 corresponding to each item response option being on a 5-point Likert scale ranging from *Strongly disagree* (1) to *Strongly agree* (5). Cronbach’s alpha (?) was used to assess internal consistency across items for each composite.^61^ We used principal components estimated with polychoric correlations to assess unidimensionality amongst items (**Supplemental Tables 4-6)**. Parallel analysis using 1000 simulated datasets was implemented to confirm construct dimensionality (**Supplemental Figures 2-3**). We used Wilcoxon signed rank tests to assess paired differences in early versus delayed action composite scores among respondents. Internal consistency and dimensionality assessment for these measures was conducted in R (v4.0.3) using the *psych* package (v2.1.9).

### Latent class analysis

Latent class analysis (LCA) was used as the preferred analytic approach in this study due to its robust and model-based statistical properties.^30,62^ Given the sample size (<500), number of initial indicators (18), and multiple response options, we employed multiple methods, including item recoding,^58^ optimized variable selection,^63,64^ and individual item inspection^62^ to improve model identification and limit boundary solution and model convergence issues. Theoretical perspectives such as respondent subgroup plausibility and interpretation, model parsimony, and model fit statistics were also considered throughout the item selection and model building process.^57^

Initially, 18 items related to perceived utility, perceived barriers, perceived benefits, and confidence in using PRS were considered as potential indicators for LCA modeling for their potential to identify subgroups of primary care physicians (PCPs) with different overall perceptions of PRS. Correlations for all items are presented in **Supplemental Table 7**.

Disease-specific utility questions and vendor-provided and self-reported demographic information were excluded from LCA modeling as potential covariates or as likely uninformative for the delineation of predicted class memberships. Raw survey response were recorded using 5-point Likert-type response categories ranging from *Strongly disagree* (1) to *Strongly agree* (5) for perceived utility, benefit, and confidence items and using 4-point Likert-type response categories ranging from *Not a barrier* (1) to *Extreme barrier* (4) for the perceived barrier items. Given the sample size and large number of items and response categories,^65^ we collapsed response data into larger categories^58^ and identified a limited number of items that yielded optimal model fit^63^ and class separation.^57^ This approach was employed to limit sparseness across contingency table cells and to ultimately improve model identification, parameter estimation, model selection, and eventual model interpretation.^30,58^

Attempts to estimate LCA models including original response categories with no covariates resulted in empirically underidentified models (specifically when the number of classes were greater than two), a large number of boundary solutions (item-response probabilities equal to 0 or 1), or unstable or failed model convergence (global, local, or flat optima differentially identified depending on start values and/or optimization algorithms used). To address these issues, we first iteratively collapsed original response options into a smaller number of categories, using the common practice of response category recoding to increase cell size in the presence of sparseness or to enhance model interpretability.^30,58,66^ Second, we used item selection techniques proposed by Fop and Murphy^63^ to further reduce data dimensionality and model complexity. Such methods can be used to exclude duplicative or uninformative variables in the spirit of parsimony^67^ and generally rely on differences in the approximations of the Bayesian information criterion (BIC) among varying models to identify items most informative to the underlying latent construct.^64,68,69^

Data were iteratively recoded considering cell size and interpretable breakpoints (e.g. condense *Strongly Disagree* and *Disagree* into single category) and potential models were re-estimated until stable solutions over a minimum of one to five classes was achieved. Consistent convergence was only achieved once items were fully dichotomized. Dichotomized items were further assessed using variable selection algorithms to remove redundant or uninformative features to further diminish the total number of response patterns. Additionally, resultant item-response probabilities were manually inspected to assess contributions to class separation. Item-response probabilities largely concentrated above or below 0.50 and with limited range between the highest and lowest probabilities (∼ < 0.40) or comparable items with redundant distributions were also considered for removal from the final indicator set^57,62^ (**Supplemental Table 8**). Model fittings across all combinations of item sets yielded generally similar class prevalences. Final item selection balanced a parsimonious model structure with both item and class interpretability. The final indicator set included seven items: four items associated with perceived barriers (“Using genetic risk scores in clinical decision-making might result in unintended harms if patients receive unnecessary testing or treatments”; “I have insufficient understanding about how to use genetic risk scores to make medical decisions”; “Genetic risk scores have been less extensively studied in non-European populations”; “I have insufficient time to explain genetic risk scores and their strengths and limitations to patients”) coded dichotomously as *Not a barrier* or *Somewhat of a barrier* versus *Moderate barrier* or *Extreme barrier*; 2 items associated with perceived benefits (“Genetic risk scores could help improve my medical decision-making in the care of patients”; “Genetic risk scores could improve my patients’ health outcomes”; and a single item related to PCP confidence (“I am confident in my ability to use genetic risk score results”, coded dichotomously as *Strongly Disagree, Disagree*, or *Neither Agree nor Disagree* versus *Agree* or *Strongly Agree*. Correlations for items included in the final indicator set are presented in **Supplemental Table 9**.

Models including one to five classes based on the final indicator set were estimated using 100 random start values. Optimal model selection was based on a variety of model fit indices, namely low BIC, a significant Bootstrapped Likelihood Ratio Test (BLRT), satisfactory relative entropy, and high average class posterior probabilities as well as considerations of class prevalences and interpretable solutions.^30,57^ All latent class modeling was conducted in R (v4.0.3) using the *mirt* (v1.35.1), *poLCA* (v1.4.1), *randomLCA* (v1.1.1), and *LCAvarsel* (v1.1) packages and in Stata (v17.0) using the *Generalized Structural Equation Model Estimatio*n (gsem) command.

## SUPPLEMENTAL RESULTS

### Latent class analysis: Model fit and interpretation

Latent class analysis supported the use of a 3-class model. **Supplemental Table 10** displays fit indices for one- to five-class solutions including our final indicator set. As expected, model complexity increased and absolute model fit (G^2^, log-likelihood) improved with a greater number of estimated classes. Relative fit indices BIC and Akaike’s Information Criterion (AIC) and Sample-size Adjusted BIC (SABIC) were minimized in the 3- and 4-class solutions, respectively. However, upon visual inspection (**Supplemental Figure 4**), AIC and SABIC differences between the 3- and 4-class solutions were determined negligible (ΔAIC=-8.087; ΔSABIC=-2.225). Likelihood ratio tests (LRT) as well as bootstrapped likelihood ratio tests (BLRT) among nested models suggested increasingly better model fits up to the 4-class solution. Classification certainty was highest (relative entropy=0.816) in the 5-class solution, but was not consequentially better than either the 3- or 4-class models (0.793 and 0.795, respectively). Additionally, average posterior probabilities indicated good class separation, which were only slightly higher in the 3-class model (range 0.886, 0.958) compared to the 4-class model (range 0.859, 0.921). Random start values were replicated (to the thousandths place) at acceptable thresholds in both the 3- (16%) and 4-class (9%) solutions, suggesting global minima were likely reached in each case.^57^ Despite relatively similar properties, we opted to move forward with the 3-class model given its lesser complexity and ease of interpretation. **Supplemental Figure 5** summarizes general interpretations, posterior probabilities, and predicted class memberships of all survey respondents (n=367) considering the final 3-class model.

