## Supplemental tables and figures for "Benefits and barriers to implementing precision preventive care: results of a national physician survey"

**SUPPLEMENTAL TABLE 1: Characteristics of physician survey non-invitees and invitees**

**SUPPLEMENTAL TABLE 2: Early action and delayed action composite score assessment, estimates, and comparison**

**SUPPLEMENTAL FIGURE 1: Disease-specific interest in polygenic risk scores**

**SUPPLEMENTAL TABLE 3: Latent class model: Item-response probabilities for 3-class solution**

**SUPPLEMENTAL TABLE 4: Correlation among responses about earlier or more intensive clinical action for high-risk polygenic risk scores**

**SUPPLEMENTAL TABLE 5: Correlation among responses about delayed or discontinued clinical action for low-risk polygenic risk scores**

**SUPPLEMENTAL TABLE 6: Eigenvalues and proportion of variance explained of early and delayed clinical action from principal component analysis**

**SUPPLEMENTAL FIGURE 2. Parallel analysis scree plot for early action items (1000 simulations)**

**SUPPLEMENTAL FIGURE 3: Parallel analysis scree plot for delayed action items (1000 simulations)**

**SUPPLEMENTAL TABLE 7: Correlation matrix for survey responses**

**SUPPLEMENTAL TABLE 8: Item selection steps for latent class analysis modeling using dichotomized items.**

**SUPPLEMENTAL TABLE 9: Correlation matrix for dichotomized items included in latent class analysis model**

**SUPPLEMENTAL TABLE 10: Fit statistics for one- to five-class solutions for latent class modeling**

**SUPPLEMENTAL FIGURE 4. Elbow plot of information criteria for 1- to 5-class solutions in latent class modeling**

**SUPPLEMENTAL FIGURE 5: General perceptions, posterior probabilities, and predicted class membership for all primary care provider respondents (n=367)**

**SUPPLEMENTAL TABLE 1: Characteristics of physician survey non-invitees and invitees**

| | **Non-invitees (*n*=235,226)** | **Non-respondents (*n*=26,631)** | **Respondents (*n*=367)** |
|---|---|---|---|
| **Mean (SD) age, years** | 50.1 (14.1) | 52.5 (13.7) | 54.9 (12.9) |
| **Mean (SD) time since medical school graduation, years** | 22.7 (14.2) | 24.6 (13.8) | 27.2 (13.3) |
| **Gender, *n* (%)** | | | |
| Female | 98,091 (41.7%)* | 10,662 (40.0%) | 137 (37.3%) |
| Male | 137,129 (58.3%)* | 15,969 (60.0%) | 230 (62.7%) |
| **Specialty, *n* (%)** | | | |
| Internal medicine | 121,072 (51.5%) | 13,497 (50.7%) | 200 (54.5%) |
| Family medicine | 108,999 (46.3%) | 12,472 (46.8%) | 159 (43.3%) |
| General practice | 5,155 (2.2%) | 662 (2.5%) | 8 (2.2%) |
| **US region, *n* (%)** | | | |
| Midwest | 52,758 (22.4%) | 5,742 (21.6%) | 82 (22.3%) |
| Northeast | 47,201 (20.0%) | 4,976 (18.7%) | 83 (22.6%) |
| South | 78,693 (33.5%) | 9,405 (35.3%) | 97 (26.4%) |
| West | 56,574 (24.1%) | 6,508 (24.4%) | 105 (28.6%) |

Non-invitees are defined as eligible primary care physicians in the IQVIA *ONEKEY* physician database who were not selected to receive the survey invitation. Non-respondents are defined as physicians who received the email invitation but did not complete at least Q1-Q6 (patient case scenarios). Respondents are defined here as respondents completing Q1-Q13. *Six non-invitees are categorized as gender “unknown” in the database.

### SUPPLEMENTAL TABLE 2: Early action and delayed action composite score assessment, estimates, and comparison

| | Internal consistency | Sampling adequacy | PCA | Score descriptives | Paired comparison |
|---|---|---|---|---|---|
| *Early and delayed item composite score internal consistency, structure, and comparison*[1] | **α (95% CI)**[2,3] | **KMO Index**[4] | **First PC Eigenvalue; % Variance Explained**[5] | **Median; (MAD)**[6] | **Wilcoxon signed rank test**[7] |
| ***In my clinical practice, I would use genetic risk scores to…."*** | | | | | |
| **Early action**<br>1. Identify high-risk patients who might need earlier or more intensive disease screening procedures.<br>2. Identify high-risk patients who might need earlier or more intensive recommendations for preventive medications.<br>3. Identify high-risk patients who might need earlier or more intensive recommendations for lifestyle modification. | 0.828 (0.765, 0.872) | 0.727 | 2.552; 0.851 | 14 (1.483) | **Test statistic; *p*-value**<br>V = 34990; $p < 0.001$<br>**Median δ (95% CI)**<br>4 (3.500, 4.500)<br>**r (95% CI)**<br>0.726 (0.699, 0.752) |
| **Delayed action**<br>1. Identify low-risk patients who might be able to delay or discontinue disease screening procedures.<br>2. Identify low-risk patients who might be able to delay or discontinue preventive medications.<br>3. Identify low-risk patients who might be able to delay or discontinue recommendations for lifestyle modification. | 0.758 (0.703, 0.803) | 0.676 | 2.214; 0.738 | 10 (2.966) | |

[1] Composite score constructed using raw Likert item data; 1 (Strongly Disagree) to 5 (Strongly Agree).
[2] Cronbach α values ≥ 0.70 are considered acceptable for internal consistency across items.
[3] Confidence intervals based on 1000 bootstrapped samples; 2.5th and 97.5th percentiles.
[4] Kaiser-Meyer-Olkin (KMO) Index; Calculated using polychoric correlations; Values ≥ 0.70 suggest data is appropriate for structural modeling.
[5] Principal components (PC) estimated using polychoric correlations; Only first PC eigenvalue > 1.000 and proportion of variance explained ≥ 0.70 suggests composite score item unidimensionality.
[6] Median and Median Absolute Deviation (MAD) calculated by summing all early action and delayed action items; composite score range = (3, 15).
[7] Paired comparison across same individuals endorsing both early and delayed action. V statistic (sum of positive scores), Median δ, and r (effect size; Z/√N) calculated by considering early action as reference composite score.
Abbreviations: PCA; Principal component analysis.

**SUPPLEMENTAL FIGURE 1: Disease-specific interest in polygenic risk scores**

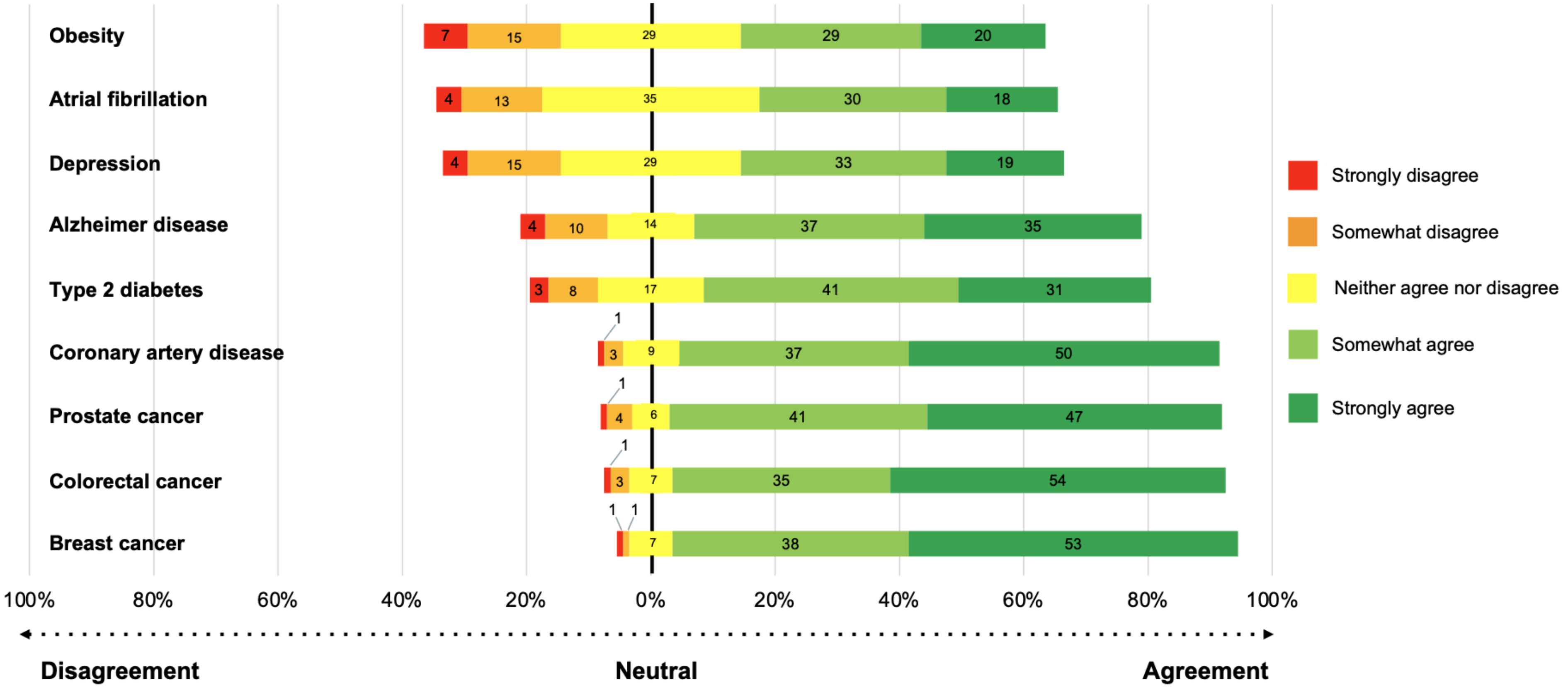


Numbers are proportions of respondents endorsing agreement or disagreement for each disease with the statement "I would be interested in using a genetic risk score test for ____________________ in my clinical practice."

**SUPPLEMENTAL TABLE 3: Latent class model: Item-response probabilities for 3-class solution**

| | Classes | | |
|---|---|---|---|
| | ***Skeptics*** | ***Learners*** | ***Enthusiasts*** |
| *Item-response probabilities (95% CI)*[1] | *Probability of endorsing moderate or extreme barrier / agree or strongly agree* | | |
| ***Barriers*** | | | |
| Using genetic risk scores in clinical decision-making might result in unintended harms if patients receive unnecessary testing or treatments. | **0.799 (0.680, 0.882)** | **0.759 (0.654, 0.840)** | 0.317 (0.238, 0.407) |
| I have insufficient understanding about how to use genetic risk scores to make medical decisions. | **0.608 (0.480, 0.723)** | **0.987 (0.889, 1.000)** | 0.099 (0.034, 0.253) |
| Genetic risk scores have been less extensively studied in non-European populations. | **0.790 (0.666, 0.876)** | **0.677 (0.573, 0.766)** | 0.273 (0.201, 0.358) |
| I have insufficient time to explain genetic risk scores and their strengths and limitations to patients. | 0.445 (0.331, 0.565) | **0.554 (0.445, 0.658)** | 0.135 (0.085, 0.209) |
| ***General measures*** | | | |
| Genetic risk scores could help improve my medical decision-making in the care of patients. | **0.585 (0.463, 0.698)** | **0.977 (0.885, 0.996)** | **0.978 (0.910, 0.995)** |
| Genetic risk scores could improve my patients' health outcomes. | 0.148 (0.041, 0.410) | **0.999 (0.999, 1.000)** | **0.908 (0.830, 0.952)** |
| I am confident in my ability to use genetic risk score results. | 0.084 (0.036, 0.183) | 0.329 (0.241, 0.432) | **0.670 (0.577, 0.752)** |

[1]Item response probabilities based on final reduced model.

Notes: Item-response probabilities > 0.50 bolded to assist interpretation. Probability of non-endorsement can be calculated by subtracting probability from 1. Confidence intervals truncated at 0.000 or 1.000. Class homogeneity is considered well-defined when conditional item probabilities are > 0.70 or < 0.30 (Nylund-Gibson & Choi, 2018).

**SUPPLEMENTAL TABLE 4: Correlation among responses about earlier or more intensive clinical action for high-risk polygenic risk scores**

| **Polychoric correlation matrix for early action items** | **1** | **2** | **3** |
|---|---|---|---|
| **1** Identify high-risk patients who might need earlier or more intensive disease screening procedures. | 1.000 | | |
| **2** Identify high-risk patients who might need earlier or more intensive recommendations for preventive medications. | **0.857** | 1.000 | |
| **3** Identify high-risk patients who might need earlier or more intensive recommendations for lifestyle modification. | **0.712** | **0.757** | 1.000 |
| **Composite score pattern matrix (unrotated loadings) from principal component analysis** | **Pattern matrix** | | |
| **Early action** | **PC1** | **PC2** | **PC3** |
| **1** Identify high-risk patients who might need earlier or more intensive disease screening procedures. | 0.931 | -0.278 | 0.239 |
| **2** Identify high-risk patients who might need earlier or more intensive recommendations for preventive medications. | 0.947 | -0.155 | -0.281 |
| **3** Identify high-risk patients who might need earlier or more intensive recommendations for lifestyle modification. | 0.889 | 0.456 | 0.050 |

Correlations ≥ |0.30| are bolded for interpretation. Principal components (PC) estimated using polychoric correlations.

**SUPPLEMENTAL TABLE 5: Correlation among responses about delayed or discontinued clinical action for low-risk polygenic risk scores**

| **Polychoric correlation matrix for delayed action items** | **1** | **2** | **3** |
|---|---|---|---|
| **1** Identify low-risk patients who might be able to delay or discontinue disease screening procedures. | 1.000 | | |
| **2** Identify low-risk patients who might be able to delay or discontinue preventive medications. | **0.751** | 1.000 | |
| **3** Identify low-risk patients who might be able to delay or discontinue recommendations for lifestyle modification. | **0.522** | **0.538** | 1.000 |
| **Composite score pattern matrix (unrotated loadings) from principal component analysis** | **Pattern matrix** | | |
| **Delayed action** | **PC1** | **PC2** | **PC3** |
| **1** Identify low-risk patients who might be able to delay or discontinue disease screening procedures. | 0.892 | -0.290 | 0.347 |
| **2** Identify low-risk patients who might be able to delay or discontinue preventive medications. | 0.899 | -0.255 | -0.357 |
| **3** Identify low-risk patients who might be able to delay or discontinue recommendations for lifestyle modification. | 0.782 | 0.624 | 0.010 |

Correlations ≥ |0.30| are bolded for interpretation. Principal components (PC) estimated using polychoric correlations.

**SUPPLEMENTAL TABLE 6: Eigenvalues and proportion of variance explained of early and delayed clinical action from principal component analysis**

| | Early action | | | Delayed action | | |
|---|---|---|---|---|---|---|
| | **PC1** | **PC2** | **PC3** | **PC1** | **PC2** | **PC3** |
| **Eigenvalues[1]** | **2.553** | 0.309 | 0.139 | **2.214** | 0.538 | 0.248 |
| **Proportion of variance explained by principal components** | | | | | | |
| Component variance | 0.851 | 0.103 | 0.046 | 0.738 | 0.179 | 0.083 |
| Cumulative variance | 0.851 | 0.954 | 1.000 | 0.738 | 0.917 | 1.000 |

[1]Eigenvalues > 1.000 represent viable components. Values > 1.000 bolded for interpretation.
Principal components (PC) estimated using polychoric correlations.

**SUPPLEMENTAL FIGURE 2. Parallel analysis scree plot for early action items (1000 simulations)**

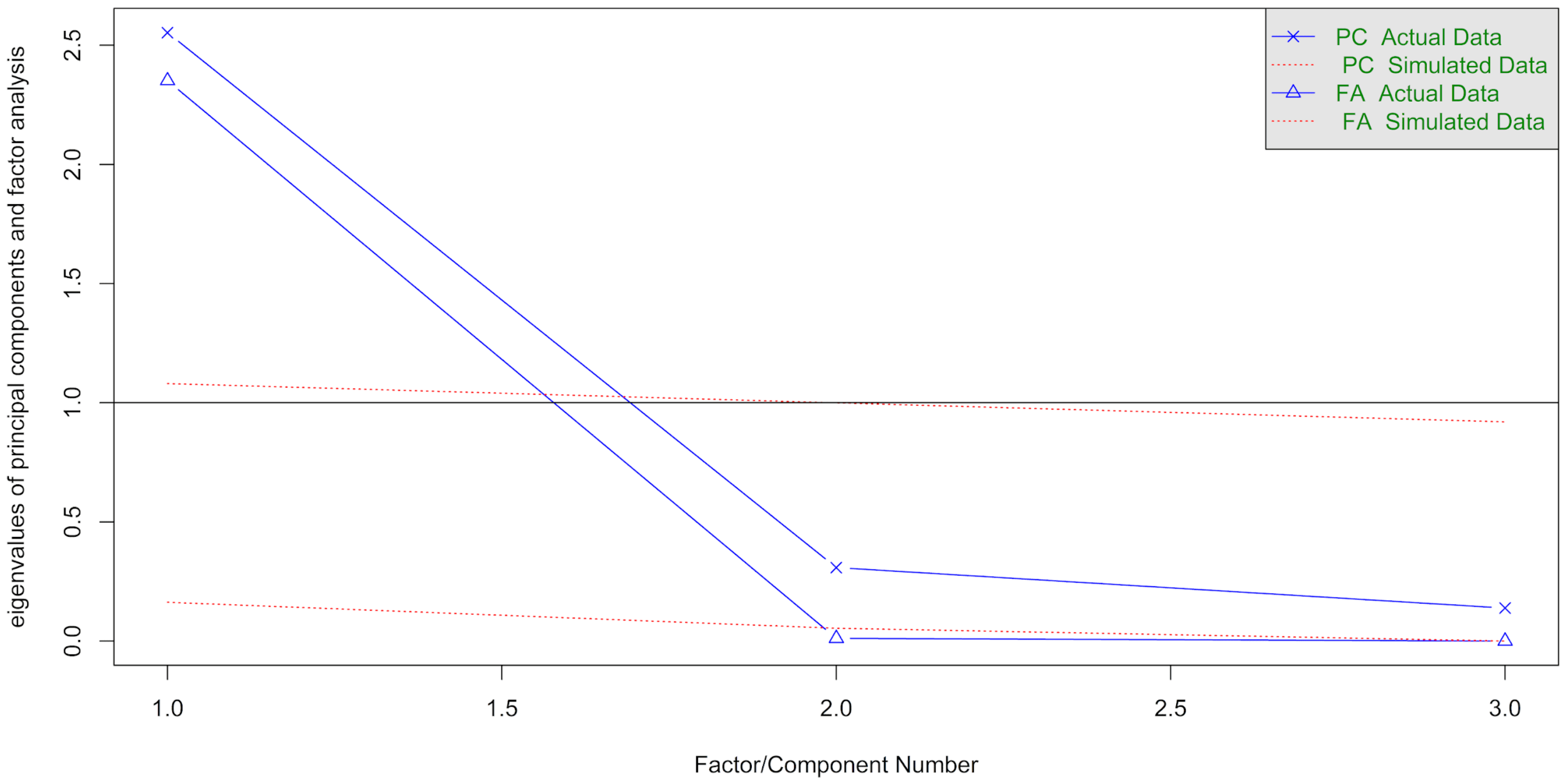

**SUPPLEMENTAL FIGURE 3: Parallel analysis scree plot for delayed action items (1000 simulations)**

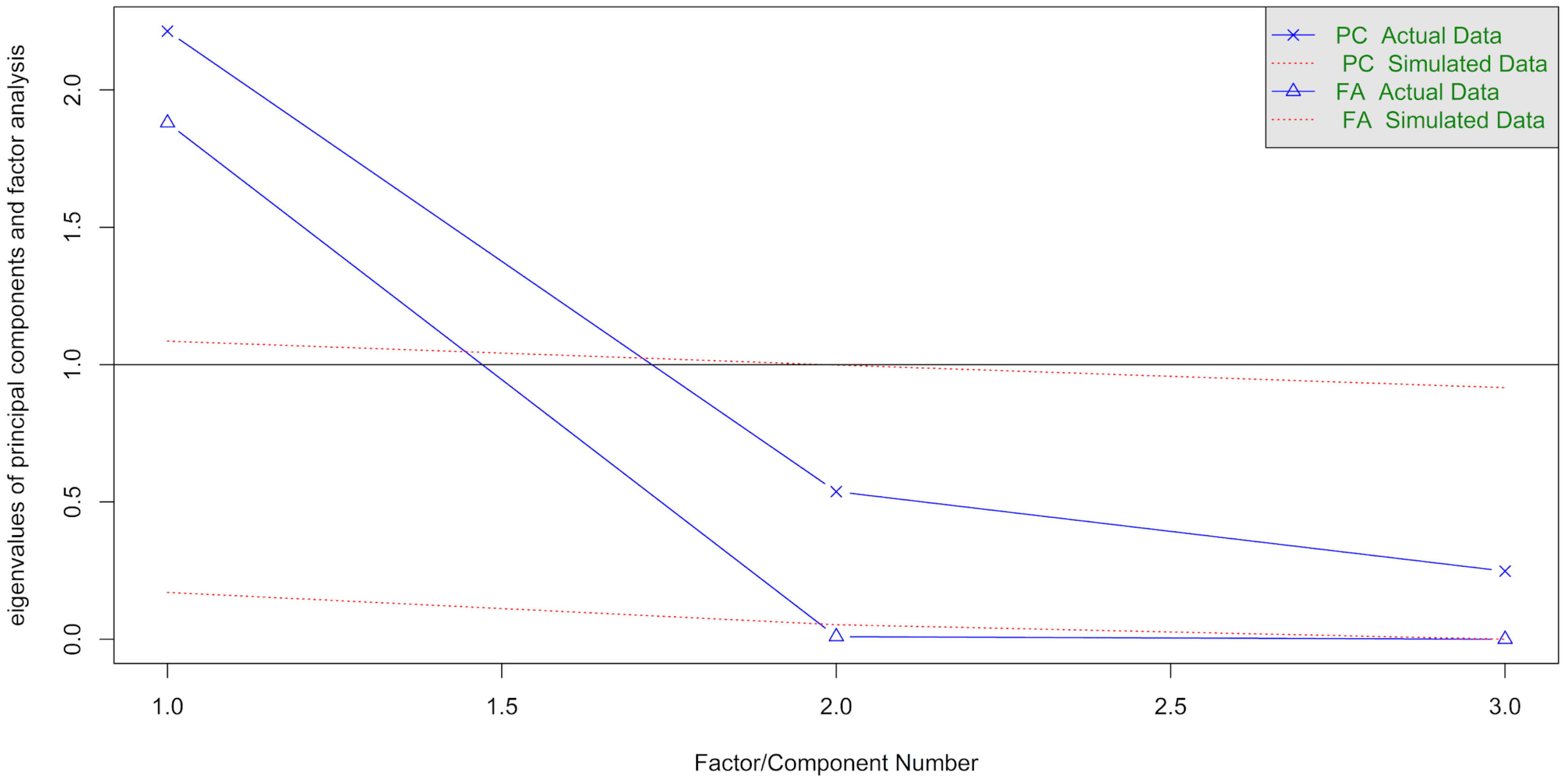

### SUPPLEMENTAL TABLE 7: Correlation matrix for survey responses

| | | 1 | 2 | 3 | 4 | 5 | 6 | 7 | 8 | 9 | 10 | 11 | 12 | 13 | 14 | 15 | 16 | 17 | 18 |
|---|---|---|---|---|---|---|---|---|---|---|---|---|---|---|---|---|---|---|---|
| *Perceived utility items* | | | | | | | | | | | | | | | | | | | |
| **1** | **Early screening** | 1.000 | | | | | | | | | | | | | | | | | |
| **2** | **Early medication** | **0.703** | 1.000 | | | | | | | | | | | | | | | | |
| **3** | **Early modification** | **0.605** | **0.628** | 1.000 | | | | | | | | | | | | | | | |
| **4** | **Delayed screening** | **0.339** | **0.340** | 0.274 | 1.000 | | | | | | | | | | | | | | |
| **5** | **Delayed medication** | **0.324** | **0.311** | **0.323** | **0.660** | 1.000 | | | | | | | | | | | | | |
| **6** | **Delayed modification** | 0.080 | 0.139 | 0.145 | **0.402** | **0.414** | 1.000 | | | | | | | | | | | | |
| *Perceived benefit items* | | | | | | | | | | | | | | | | | | | |
| **7** | **Decision-making** | **0.495** | **0.526** | **0.371** | 0.287 | 0.260 | 0.137 | 1.000 | | | | | | | | | | | |
| **8** | **Patient decision-making** | **0.406** | **0.433** | **0.354** | 0.239 | 0.208 | 0.129 | **0.733** | 1.000 | | | | | | | | | | |
| **9** | **Health outcomes** | **0.314** | **0.370** | 0.264 | 0.252 | 0.201 | 0.128 | **0.628** | **0.603** | 1.000 | | | | | | | | | |
| *Perceived confidence* | | | | | | | | | | | | | | | | | | | |
| **10** | **Confidence** | 0.208 | 0.224 | 0.220 | 0.198 | 0.176 | 0.161 | **0.332** | 0.285 | **0.345** | 1.000 | | | | | | | | |
| *Perceived barrier items* | | | | | | | | | | | | | | | | | | | |
| **11** | **Cost** | -0.007 | -0.017 | -0.055 | -0.027 | -0.088 | -0.147 | -0.052 | -0.012 | -0.037 | -0.181 | 1.000 | | | | | | | |
| **12** | **Anxiety** | -0.128 | -0.095 | -0.050 | -0.037 | -0.016 | -0.062 | -0.090 | -0.069 | -0.068 | -0.164 | 0.207 | 1.000 | | | | | | |
| **13** | **Discrimination** | -0.133 | -0.086 | -0.033 | -0.038 | -0.015 | -0.015 | -0.046 | -0.017 | -0.067 | -0.084 | 0.206 | **0.343** | 1.000 | | | | | |
| **14** | **Unnecessary treatment** | -0.224 | -0.173 | -0.166 | -0.020 | -0.080 | 0.020 | -0.176 | -0.114 | -0.176 | -0.258 | 0.141 | **0.313** | 0.271 | 1.000 | | | | |
| **15** | **No guidelines** | -0.209 | -0.140 | -0.177 | -0.104 | -0.104 | -0.088 | -0.163 | -0.108 | -0.157 | -0.273 | 0.215 | 0.228 | 0.165 | **0.374** | 1.000 | | | |
| **16** | **Provider understanding** | -0.135 | -0.119 | -0.110 | -0.054 | -0.052 | -0.003 | -0.149 | -0.088 | -0.087 | **-0.398** | 0.187 | 0.203 | 0.085 | **0.324** | **0.426** | 1.000 | | |
| **17** | **Non-European population** | -0.174 | -0.122 | -0.095 | -0.063 | -0.029 | -0.069 | -0.163 | -0.165 | -0.176 | -0.226 | 0.112 | 0.215 | 0.169 | 0.250 | **0.385** | **0.339** | 1.000 | |
| **18** | **Time** | -0.205 | -0.188 | -0.156 | -0.032 | -0.083 | -0.017 | -0.186 | -0.113 | -0.111 | -0.206 | 0.016 | 0.217 | 0.153 | 0.225 | 0.158 | 0.282 | 0.218 | 1.000 |

Correlation coefficients computed as Kendall's Tau (τ). Correlations ≥ |0.30| are bolded for interpretation.

**SUPPLEMENTAL TABLE 8: Item selection steps for latent class analysis modeling using dichotomized items.**

| Candidate items | | Stepwise backward selection[1] | Swap-stepwise backward selection[2] | For manual inspection[3] | Assessment decision upon manual inspection[4,5] | Final item set |
|---|---|---|---|---|---|---|
| *Perceived utility items* | | | | | | |
| 1 | **Early screening** | **Retain** | **Retain** | **Retain** | Drop; Low class separation | Drop |
| 2 | **Early medication** | **Retain** | **Retain** | **Retain** | Drop; Low class separation | Drop |
| 3 | **Early modification** | **Retain** | Drop | **Retain** | Drop; Low class separation | Drop |
| 4 | **Delayed screening** | Drop | Drop | Drop | | Drop |
| 5 | **Delayed medication** | Drop | Drop | Drop | | Drop |
| 6 | **Delayed modification** | Drop | Drop | Drop | | Drop |
| *Perceived benefit/confidence items* | | | | | | |
| 7 | **Decision-making** | **Retain** | **Retain** | **Retain** | Retain | **Retain** |
| 8 | **Patient decision-making** | **Retain** | **Retain** | **Retain** | Drop; Redundant item-probabilities | Drop |
| 9 | **Health outcomes** | **Retain** | **Retain** | **Retain** | Retain | **Retain** |
| 10 | **Confidence** | **Retain** | **Retain** | **Retain** | Retain | **Retain** |
| *Perceived barrier items* | | | | | | |
| 11 | **Cost** | Drop | Drop | Drop | | Drop |
| 12 | **Anxiety** | Drop | Drop | Drop | | Drop |
| 13 | **Discrimination** | Drop | Drop | Drop | | Drop |
| 14 | **Unnecessary treatment** | **Retain** | **Retain** | **Retain** | Retain | **Retain** |
| 15 | **No guidelines** | Drop | Drop | Drop | | Drop |
| 16 | **Provider understanding** | **Retain** | **Retain** | **Retain** | Retain | **Retain** |
| 17 | **Non-European population** | **Retain** | **Retain** | **Retain** | Retain | **Retain** |
| 18 | **Time** | **Retain** | **Retain** | **Retain** | Retain | **Retain** |

Two- to five- class model solutions fit over each item set using 50 random start values. All potentially meaningful items retained for consideration in final item set.
All item assessment undertaken with fully dichotomized items: Strongly Disagree, Disagree, Neither Agree nor Disagree versus Agree, Strongly Agree; Not a barrier, Somewhat of a barrier versus Moderate barrier, Extreme barrier.
[1,2]Item selection via the stepwise and swap-stepwise selection algorithms proposed by Fop, Smart, & Murphy, 2017 and Fop & Murphy, 2018.
[1]The algorithm starts with a model including all variables, at each step a variable is removed or added and competing model configurations are compared using differences in approximated BIC.
[2]The algorithm starts with a model including all variables, at each step a variable is removed or added and competing model configurations are compared using differences in approximated BIC. An additional swap step occurs at each iteration comparing two separate model configurations, one including the current model configuration and a second including a non-clustering variable.
[3]Only variables retained by algorithms were considered for further inspection.
[4]Perceived utility items demonstrated low class separation (concentrated item-response probabilities above 0.50) and/or contributed to small class prevalences (<0.04) in 4- and 5- class models.
[5]Patient decision-making item-response probabilities were largely redundant with physician decision-making. Physician decision-making retained due to theoretical considerations.

**SUPPLEMENTAL TABLE 9: Correlation matrix for dichotomized items included in latent class analysis model**

| | 1 | 2 | 3 | 4 | 5 | 6 | 7 |
|---|---|---|---|---|---|---|---|
| *Perceived benefit items* | | | | | | | |
| **1 Decision-making** | 1.000 | | | | | | |
| **2 Health outcomes** | **0.416** | 1.000 | | | | | |
| *Perceived confidence* | | | | | | | |
| **3 Confidence** | 0.163 | **0.304** | 1.000 | | | | |
| *Perceived barrier items* | | | | | | | |
| **4 Unnecessary treatment** | -0.165 | -0.150 | -0.202 | 1.000 | | | |
| **5 Provider understanding** | -0.052 | -0.012 | -0.269 | **0.352** | 1.000 | | |
| **6 Non-European population** | -0.164 | -0.190 | -0.171 | 0.210 | **0.351** | 1.000 | |
| **7 Time** | -0.124 | -0.030 | -0.170 | 0.197 | **0.320** | 0.209 | 1.000 |

Correlation coefficients computed as Kendall's Tau (τ) (equivalent to phi coefficient (ϕ)). Correlations ≥ |0.30| are bolded for interpretation.

**SUPPLEMENTAL TABLE 10: Fit statistics for one- to five-class solutions for latent class modeling**

| Number of latent classes estimated | Number of parameters estimated | $G^2$ | AIC | BIC | SABIC | LL | LRT (*p*) | BLRT[1] (*p*) | Entropy[2] |
|---|---|---|---|---|---|---|---|---|---|
| 1 | 7 | 397.782 | 3160.716 | 3188.054 | 3165.846 | -1573.358 | - | - | - |
| 2 | 15 | 188.441 | 2967.374 | 3025.955 | 2978.366 | -1468.687 | < 0.001 | < 0.001 | 0.661 |
| 3 | 23 | 108.086 | 2903.019 | **2992.843** | 2919.872 | -1428.510 | < 0.001 | < 0.001 | 0.793 |
| 4 | 31 | 83.999 | **2894.932** | 3015.999 | **2917.647** | -1416.466 | **0.002** | **0.015** | 0.795 |
| 5 | 39 | **71.686** | 2898.619 | 3050.928 | 2927.196 | **-1410.310** | 0.138 | 0.522 | **0.816** |

Note: All models estimated using 100 random start values. Multiple fit statistics reported, model selection involved fit assessment, interpretability, parsimony, and theoretical perspectives (Weller et al., 2020). Best relative fit indices bolded for interpretation.

[1] p-value generated using 1000 samples.

[2] Entropy ≥ 0.80 demonstrates reliable class assignment (Clark & Muthén, 2009; Nylund-Gibson & Choi, 2018).

Abbreviations: AIC = Akaike information criterion; BIC = Bayesian information criterion; SABIC = Sample-size adjusted BIC; LL = log-likelihood; LRT = likelihood ratio test; BLRT = Bootstrapped likelihood ratio test.

**SUPPLEMENTAL FIGURE 4. Elbow plot of information criteria for 1- to 5-class solutions in latent class modeling**

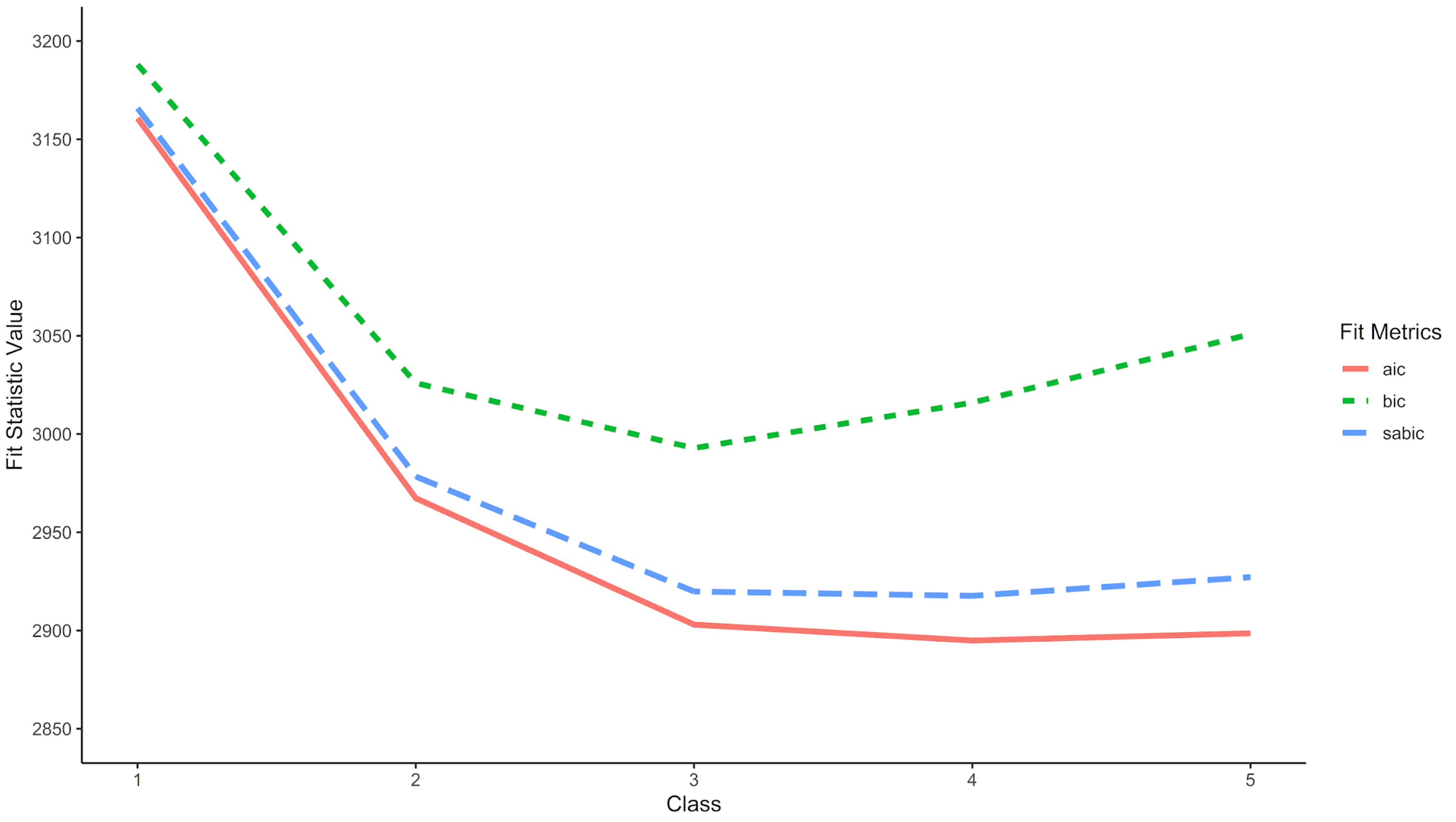

**SUPPLEMENTAL FIGURE 5: General perceptions, posterior probabilities, and predicted class membership for all primary care provider respondents (n=367)**

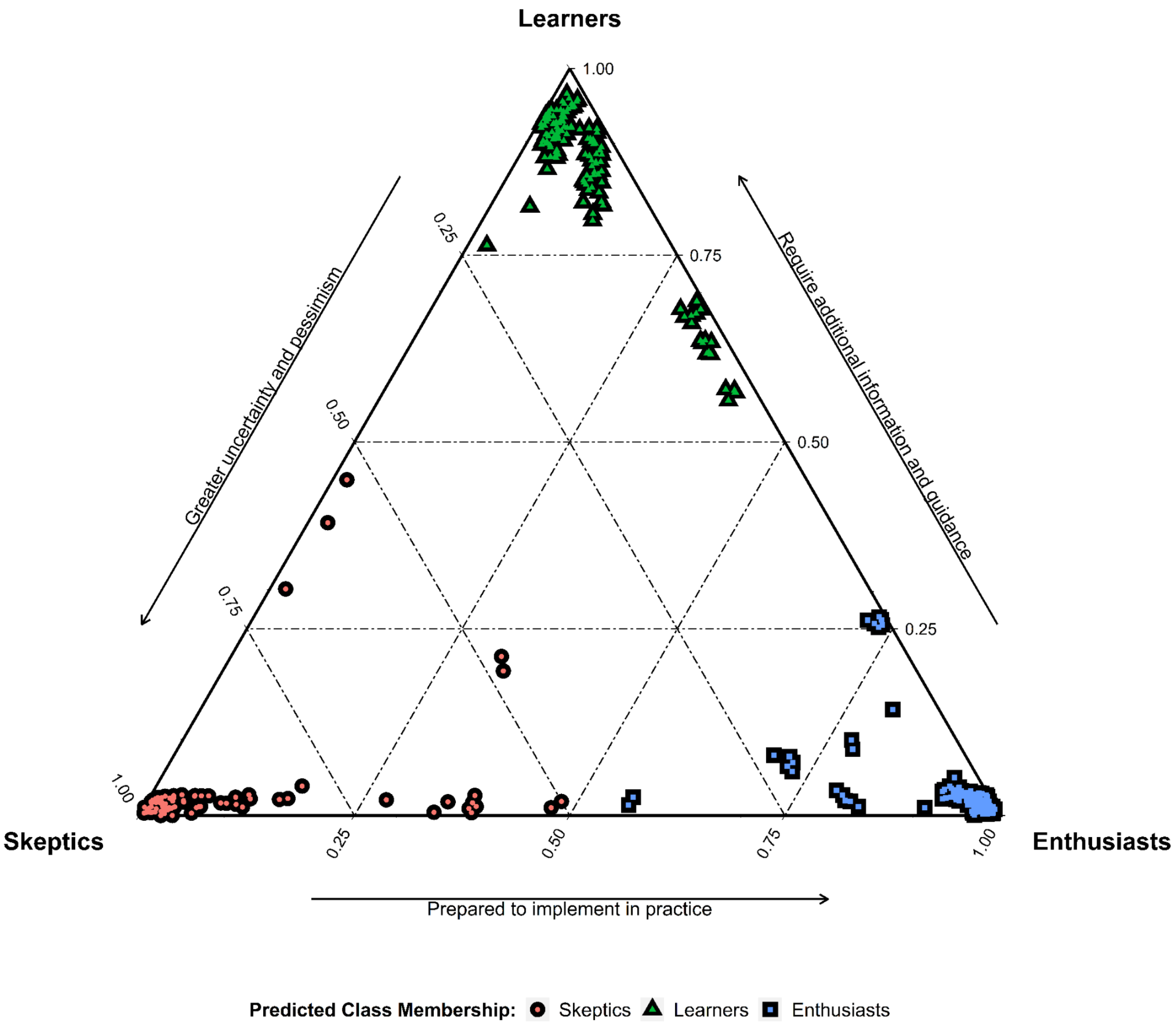
