## Supplementary material for "Benefits and barriers to implementing precision preventive care: results of a national physician survey": Survey instrument

### Survey on precision prevention in primary care

#### Genetic risk scores for disease prevention in primary care

This research is being conducted to learn about physician perceptions of the clinical utility of **genetic risk scores**, also known as polygenic risk scores. Genetic risk scores may soon be available for primary care physicians (PCPs) to order for their patients, and PCPs may increasingly be asked to understand, and make clinical recommendations based on, their patients' genetic risk score results. You are being asked to participate in this study as a primary care physician.

The survey will ask you to respond to a few patient cases, followed by a few general questions about genetic risk scores. It will take about 8-10 minutes. There are no direct benefits to you for your taking part in this research, but you will receive a $50 Amazon gift card to thank you for completion of this survey.

Your participation is voluntary and may be withdrawn at any time for any reason. Participation will have no effect on employment status and supervisors will not have access to survey responses.

If you have any questions about this study, please contact Dr. Benjamin Kerman at.

Please click "**Next**" to agree to participate in the research study and proceed to the survey.

**Please read the information below for a basic introduction to genetic risk scores.**

A **genetic risk score** is a number summarizing an individual's genetic predisposition to a certain trait or complex disease.

- Unlike genetic results for classic single-gene diseases like cystic fibrosis and sickle cell disease, a genetic risk score adds up the information from hundreds or even millions of genetic locations identified to be associated with a given disease.
- A patient's genetic risk score can place them on a bell curve of genetic susceptibility for the disease (see figure below).
- Genetic risk scores now exist for dozens of common diseases such as coronary artery disease and prostate cancer.
- Genetic risk scores do not take into account other disease risk factors, such as family history, smoking, or obesity.
- They have been most extensively studied in populations of European descent.
- Genetic risk scores may emerge as an important part of disease prediction, prevention, and screening in the future of primary care practice.

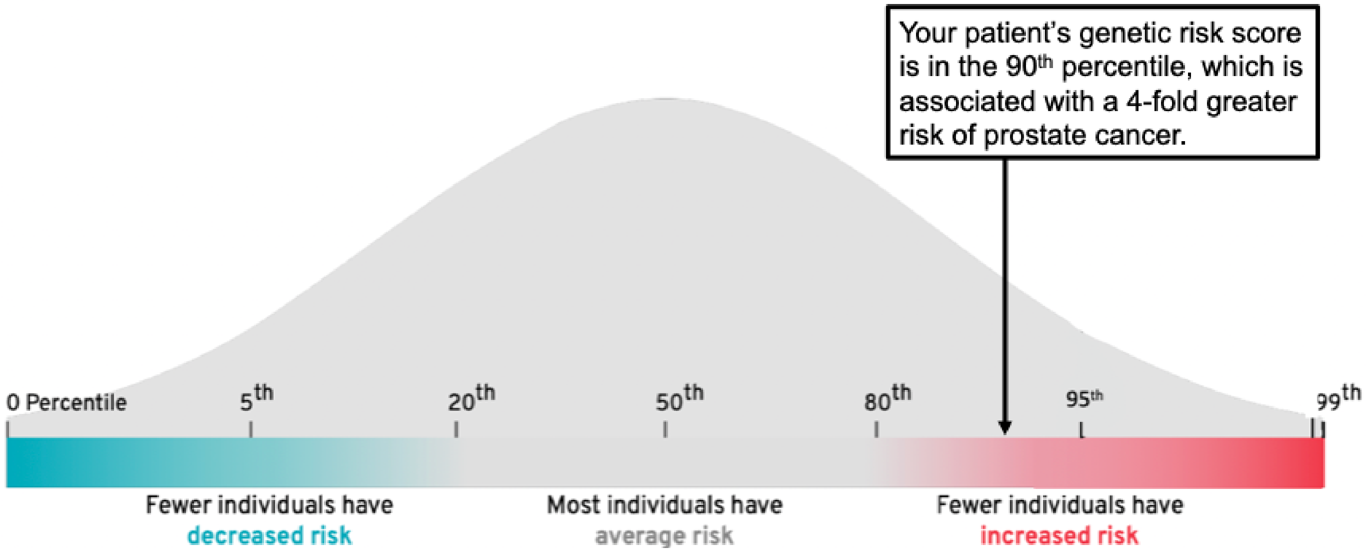


**Distribution of genetic risk scores for prostate cancer in a population, along with the results from an example patient.**

**Please click "Next" to begin the survey.** Because genetic risk scores are still a new area of research, there are no right or wrong answers to the following survey questions.

#### Case Scenarios: Coronary Artery Disease

These patient scenarios focus on the **primary prevention of atherosclerotic cardiovascular disease (ASCVD)**, including myocardial infarction and stroke, among patients at intermediate risk (10-year risk of cardiovascular event 5%-10%). Current guidelines differ on whether to initiate statin therapy for primary prevention in this group:

- **United States Preventive Services Task Force (2016)**: Recommends statin therapy for patients who: 1) are aged 40-75; 2) have 1 or more cardiovascular risk factor [dyslipidemia (LDL cholesterol >130 mg/dL), diabetes, hypertension, or smoking]; and 3) have calculated 10-year risk of cardiovascular event ≥10% (Grade B recommendation). It gives a Grade C recommendation (net benefit is small) for statin therapy in such patients with intermediate risk (10-year risk 5%-10%).
- **American College of Cardiology/American Heart Association (2019)**: Recommends statin therapy for patients ages 40-75 with LDL cholesterol between 70-190 mg/dL> who have a 10-year risk of cardiovascular event ≥7.5%.
- In your own clinical practice, your approach to statin therapy for primary ASCVD prevention might use these guidelines or other strategies, including measurement of coronary calcium scores, C-reactive protein levels, or other risk factors.

**Please click “Next” to proceed to the patient case scenarios.**

**Patient A** is a 60-year-old generally healthy man who identifies as European-American (white). He has no personal or family history of cardiovascular disease or other ASCVD risk factors. His LDL-C is 120 mg/dL, and his calculated **10-year ASCVD risk is 7.5% (intermediate risk)**. He takes no medications and reports no symptoms of coronary artery disease. He has expressed no preference for or against ASCVD prevention and asks for your recommendation.

**Please indicate how likely you are to agree with the following statements.**

**Q1. “Assuming shared decision-making, I would take the following clinical actions**

**for a patient like Patient A."**

| | Strongly DISAGREE | Somewhat DISAGREE | Neither agree nor disagree | Somewhat AGREE | Strongly AGREE |
|---|---|---|---|---|---|
| Recommend a statin | ○ | ○ | ○ | ○ | ○ |
| Order additional testing (e.g. stress test, EKG, coronary calcium score) | ○ | ○ | ○ | ○ | ○ |
| Refer to a specialist (e.g. cardiologist) | ○ | ○ | ○ | ○ | ○ |

**Patient B** has the same clinical presentation as Patient A. That is, he is a 60-year-old generally healthy man who identifies as European-American (white). He has no personal or family history of cardiovascular disease or other ASCVD risk factors. His LDL-C is 120 mg/dL, and his calculated **10-year ASCVD risk is 7.5% (intermediate risk)**. He takes no medications and reports no symptoms of coronary artery disease. In addition, you ordered a genetic risk score for coronary artery disease, which was reported as **HIGH genetic risk (90th percentile, 3-fold higher genetic risk of coronary disease compared to the average person)**. He has expressed no preference for or against ASCVD prevention and asks for your recommendation.

**Please indicate how likely you are to agree with the following statements.**

**Q2. "Assuming shared decision-making, I would take the following clinical actions for a patient like Patient B"**

| | Strongly DISAGREE | Somewhat DISAGREE | Neither agree nor disagree | Somewhat AGREE | Strongly AGREE |
|---|---|---|---|---|---|
| Recommend a statin | ○ | ○ | ○ | ○ | ○ |

| | Strongly DISAGREE | Somewhat DISAGREE | Neither agree nor disagree | Somewhat AGREE | Strongly AGREE |
|---|---|---|---|---|---|
| Order additional testing (e.g. stress test, EKG, coronary calcium score) | ○ | ○ | ○ | ○ | ○ |
| Refer to a specialist (e.g. cardiologist, geneticist) | ○ | ○ | ○ | ○ | ○ |

**Patient C** has the same clinical presentation as Patient A. That is, he is a 60-year-old generally healthy man who identifies as European-American (white). He has no personal or family history of cardiovascular disease or other ASCVD risk factors. His LDL-C is 120 mg/dL, and his calculated **10-year ASCVD risk is 7.5% (intermediate risk)**. He takes no medications and reports no symptoms of coronary artery disease. In addition, you ordered a genetic risk score for coronary artery disease, which was reported as **LOW genetic risk (10th percentile, 50% lower genetic risk of coronary disease compared to the average person)**. He has expressed no preference for or against ASCVD prevention and asks for your recommendation.

**Please indicate how likely you are to agree with the following statements.**

**Q3. "Assuming shared decision-making, I would take the following clinical actions for a patient like Patient C"**

| | Strongly DISAGREE | Somewhat DISAGREE | Neither agree nor disagree | Somewhat AGREE | Strongly AGREE |
|---|---|---|---|---|---|
| Recommend a statin | ○ | ○ | ○ | ○ | ○ |
| Order additional testing (e.g. stress test, EKG, coronary calcium score) | ○ | ○ | ○ | ○ | ○ |
| Refer to a specialist (e.g. cardiologist, geneticist) | ○ | ○ | ○ | ○ | ○ |

**Case Scenarios: Prostate Cancer**

The next clinical scenarios focus on **screening for prostate cancer.** Current guidelines do not definitively endorse prostate cancer screening for all men:

- **United States Preventive Services Task Force (2018)**: Recommends prostate cancer screening with prostate-specific antigen **(PSA) testing for men aged 55-69** only after an individualized discussion of the risks and benefits of screening (**Grade C recommendation**: net benefit is small). African-American men and men with a family history of prostate cancer are noted to be at higher risk, but the task force does not make separate screening recommendations for these groups.
- Recommendations from the **American Urological Association (2013)**, the **American College of Physicians (2013)**, and other professional organizations similarly advocate shared decision-making around PSA-based **prostate cancer screening for men aged 55-69**.
- Most guidelines do not recommend a digital rectal exam (DRE) for prostate cancer screening.
- In your own clinical practice, your approach to prostate cancer screening might use these guidelines or other strategies.

**Please click "Next" to proceed to the patient case scenarios.**

**Patient D** is a 45-year-old generally healthy European-American (white) man without personal or family history of prostate cancer. He takes no medications and reports no symptoms of prostate cancer. He has expressed no preference for or against PSA testing for prostate cancer screening and asks for your recommendation.

**Q4. Assuming shared decision-making, which prostate cancer screening strategy would you be most likely to recommend for Patient D?**

- ○ Begin PSA screening now (age 45)
- ○ Begin PSA screening at age 50
- ○ Begin PSA screening at age 55
- ○ Begin PSA screening at age 60

- Do not recommend PSA screening

**Patient E** has the same clinical presentation as Patient D. That is, he is a 45-year-old generally healthy European-American (white) man without personal or family history of prostate cancer. He takes no medications and reports no symptoms of prostate cancer. He has expressed no preference for or against PSA testing for prostate cancer screening and asks for your recommendation. **You ordered a genetic risk score for prostate cancer, which was reported as LOW genetic risk (10th percentile, 50% lower genetic risk of prostate cancer compared to the average person).**

**Q5. Assuming shared decision-making, which prostate cancer screening strategy would you be most likely to recommend for Patient E?**

- Begin PSA screening now (age 45)
- Begin PSA screening at age 50
- Begin PSA screening at age 55
- Begin PSA screening at age 60
- Do not recommend PSA screening

**Patient F** has the same clinical presentation as Patient D. That is, he is a 45-year-old generally healthy European-American (white) man without personal or family history of prostate cancer. He takes no medications and reports no symptoms of prostate cancer. He has expressed no preference for or against PSA testing for prostate cancer screening and asks for your recommendation. **You ordered a genetic risk score for prostate cancer, which was reported as HIGH genetic risk (90th percentile, ~4-fold higher genetic risk of prostate cancer compared to the average person).**

**Q6. Assuming shared decision-making, which prostate cancer screening strategy would you be most likely to recommend for Patient F?**

- Begin PSA screening now (age 45)
- Begin PSA screening at age 50
- Begin PSA screening at age 55
- Begin PSA screening at age 60

○ Do not recommend PSA screening

**GENERAL QUESTIONS**

Assume that genetic risk scores are available to estimate your patients' genetic risk for common diseases.

**Q7. Please indicate how likely you are to agree with the following statements:**

**"In my clinical practice, I would use genetic risk scores to ______________."**

| | Strongly DISAGREE | Somewhat DISAGREE | Neither agree nor disagree | Somewhat AGREE | Strongly AGREE |
|---|---|---|---|---|---|
| Identify **high-risk** patients who might need **earlier or more intensive disease screening** procedures (e.g. mammography, colonoscopy) | ○ | ○ | ○ | ○ | ○ |
| Identify **low-risk** patients who might be able to **delay or discontinue disease screening** procedures (e.g. mammography, colonoscopy) | ○ | ○ | ○ | ○ | ○ |
| Identify **high-risk** patients who might need **earlier or more intensive recommendations for preventive medications** (e.g. statins, aspirin, tamoxifen) | ○ | ○ | ○ | ○ | ○ |

| | Strongly DISAGREE | Somewhat DISAGREE | Neither agree nor disagree | Somewhat AGREE | Strongly AGREE |
|---|---|---|---|---|---|
| Identify **low-risk** patients who might be able to **delay or discontinue preventive medications** (e.g. statins, aspirin, tamoxifen) | ○ | ○ | ○ | ○ | ○ |
| Identify **high-risk** patients who might need **earlier or more intensive recommendations for lifestyle modification** (e.g. weight loss, smoking cessation) | ○ | ○ | ○ | ○ | ○ |
| Identify **low-risk** patients who might be able to **delay or discontinue recommendations for lifestyle modification** (e.g. weight loss, smoking cessation) | ○ | ○ | ○ | ○ | ○ |

**Q8. To what degree are the following items potential BARRIERS to using genetic risk scores in your clinical practice?**

| | NOT a barrier | SOMEWHAT of a barrier | MODERATE barrier | EXTREME barrier |
|---|---|---|---|---|
| Patients may incur out-of-pocket costs if insurance does not cover genetic risk score testing. | ○ | ○ | ○ | ○ |
| Patients may have anxiety about genetic risk score results. | ○ | ○ | ○ | ○ |
| Patients may have concerns about insurance discrimination. | ○ | ○ | ○ | ○ |

| | NOT a barrier | SOMEWHAT of a barrier | MODERATE barrier | EXTREME barrier |
|---|---|---|---|---|
| Using genetic risk scores in clinical decision-making might result in unintended harms if patients receive unnecessary testing or treatments. | ○ | ○ | ○ | ○ |
| There are no clinical guidelines for how to use genetic risk scores in practice (e.g. from professional organizations or my own institution). | ○ | ○ | ○ | ○ |
| I have insufficient understanding about how to use genetic risk scores to make medical decisions. | ○ | ○ | ○ | ○ |
| Genetic risk scores have been less extensively studied in non-European populations. | ○ | ○ | ○ | ○ |
| I have insufficient time to explain genetic risk scores and their strengths and limitations to patients. | ○ | ○ | ○ | ○ |

Assume that genetic risk scores are available to estimate your patients' genetic risk for the diseases in the table below.

**How strongly do you agree with this statement for each of the following diseases?**

**Q9. "I would be interested in using a genetic risk score test for ________________ in my clinical practice."**

| | Strongly DISAGREE | Somewhat DISAGREE | Neither agree nor disagree | Somewhat AGREE | Strongly AGREE |
|---|---|---|---|---|---|
| Breast cancer | ○ | ○ | ○ | ○ | ○ |

| | Strongly DISAGREE | Somewhat DISAGREE | Neither agree nor disagree | Somewhat AGREE | Strongly AGREE |
|---|---|---|---|---|---|
| Colorectal cancer | ○ | ○ | ○ | ○ | ○ |
| Prostate cancer | ○ | ○ | ○ | ○ | ○ |
| Coronary artery disease | ○ | ○ | ○ | ○ | ○ |
| Atrial fibrillation | ○ | ○ | ○ | ○ | ○ |
| Type 2 diabetes | ○ | ○ | ○ | ○ | ○ |
| Obesity | ○ | ○ | ○ | ○ | ○ |
| Alzheimer disease | ○ | ○ | ○ | ○ | ○ |
| Major depressive disorder | ○ | ○ | ○ | ○ | ○ |

**Q10. Please indicate your level of agreement with the following statements about genetic risk scores in general.**

| | Strongly DISAGREE | Somewhat DISAGREE | Neither agree nor disagree | Somewhat AGREE | Strongly AGREE |
|---|---|---|---|---|---|
| Genetic risk scores could help **improve my medical decision-making** in the care of patients. | ○ | ○ | ○ | ○ | ○ |
| Genetic risk scores could **help my patients make better decisions about their health care.** | ○ | ○ | ○ | ○ | ○ |
| Genetic risk scores could **improve my patients' health outcomes.** | ○ | ○ | ○ | ○ | ○ |
| I am **confident in my ability to use genetic risk score results.** | ○ | ○ | ○ | ○ | ○ |

**YOUR BACKGROUND**

**Q11.** “Have you received any genetics education beyond the typical medical school curriculum? (Check all that apply)

- ☐ I have had no additional formal genetic training
- ☐ Genetics residency or fellowship
- ☐ Genetics education course (online or in person)
- ☐ Residency rotation in Genetics
- ☐ Graduate degree (in addition to your professional degree) focused on genetics
- ☐ Other – Specify: ______

**Q12.** With which of the following racial groups do you identify? Please check all that apply.

- ☐ American Indian or Alaska Native
- ☐ Asian
- ☐ Black or African American
- ☐ Native Hawaiian or Other Pacific Islander
- ☐ White
- ☐ Other
- ☐ Prefer not to answer

**Q13.** Do you identify as Latinx or Hispanic?

- ○ Yes
- ○ No
- ○ Prefer not to answer
