## Supplementary material for "Benefits and barriers to implementing precision preventive care: results of a national physician survey": CHERRIES checklist

**Checklist for Reporting Results of Internet E-Surveys (CHERRIES)**

| Checklist Item | Explanation | Application to current study |
|---|---|---|
| **Category: Design** | | |
| **1. Describe survey design** | 1. Describe target population, sample frame. Is the sample a convenience sample? | The target population was primary care physicians who care for adult patients, which we defined as physicians practicing family medicine, general practice, or internal medicine. We worked with database licensee IQVIA to recruit respondents from their *ONEKEY* proprietary national physician database of from the American Medical Association (AMA) Physician Masterfile and other sources. The *ONEKEY* database contains data from more than 250,000 active physicians who have opted in to receiving email survey invitations. |
| **Category: IRB approval and informed consent process** | | |
| **1. IRB approval** | 1. Mention whether the study has been approved by an IRB. | 1. The study was approved by the Harvard Longwood Campus IRB (protocol #20-2098). |
| **2. Informed consent** | 2. Describe the informed consent process. Where were the participants told the length of time of the survey, which data were stored and where and for how long, who the investigator was, and the purpose of the study? | 2. The invitation emails described the survey as an 8-10 minute survey about precision prevention in primary care using genetic risk scores. Recipients were able to opt out of further contact using a link in the invitation email. The survey link brought physicians to a brief informed consent page, at the bottom of which respondents electronically documented consent by clicking to proceed to the first page of the survey. The respondents were told the names of the principal investigators. |
| **3. Data protection** | 3. If any personal information was collected or stored, describe what mechanisms were used to protect unauthorized access. | 3. Participant-level data, including survey responses, email addresses, and Internet Protocol addresses were stored on encrypted, password-protected servers accessible only to approved staff. |

| | | |
|---|---|---|
| **Category: Development and pre-testing** | | |
| **1. Development and testing** | 1. State how the survey was developed, including whether the usability and technical functionality of the electronic questionnaire had been tested before fielding the questionnaire. | Survey questions included novel and adapted, published instruments. The electronic survey was piloted among eight primary care practitioners for comprehension and interpretation of questions before fielding. |
| **Category: Recruitment process and description of the sample having access to the questionnaire** | | |
| **1. Open survey versus closed survey** | 1. An "open survey" is a survey open for each visitor of a site, while a closed survey is only open to a sample which the investigator knows (password-protected survey). | 1. This was a closed survey. |
| **2. Contact mode** | 2. Indicate whether or not the initial contact with the potential participants was made on the Internet. (Investigators may also send out questionnaires by mail and allow for Web-based data entry.) | 2. Initial contact with potential participants was made via email from database licensee IQVIA, which contained unique web links to the survey. |
| **3. Advertising the survey** | 3. How/where was the survey announced or advertised? Some examples are offline media (newspapers), or online (mailing lists – If yes, which ones?) or banner ads (Where were these banner ads posted and what did they look like?). It is important to know the wording of the announcement as it will heavily influence who chooses to participate. Ideally the survey announcement should be published as an appendix. | 3. The survey was introduced via email, with subject lines describing the survey as one about precision prevention in primary care. |
| **Category: Survey administration** | | |
| **1. Web/E-mail** | 1. State the type of e-survey (eg, one posted on a Web site, or one sent out through e-mail). If it is an e-mail survey, were the responses entered manually into a database, or was there an automatic method for capturing responses? | 1. This was a web-based survey accessible by email invitation only. |
| **2. Context** | 2. Describe the Web site (for mailing list/newsgroup) in which the survey was posted. What is the Web site about, who is | 2. The proprietary IQVIA *ONEKEY* national physician database of from the American Medical Association (AMA) Physician |

| | | |
|---|---|---|
| | visiting it, what are visitors normally looking for? Discuss to what degree the content of the Web site could pre-select the sample or influence the results. For example, a survey about vaccination on an anti-immunization Web site will have different results from a Web survey conducted on a government Web site. | Masterfile and other sources. The *ONEKEY* database contains data from more than 250,000 active physicians who have opted in to receiving email survey invitations. |
| **3. Mandatory/voluntary** | 3. Was it a mandatory survey to be filled in by every visitor who wanted to enter the Web site, or was it a voluntary survey? | 3. N/A |
| **4. Incentives** | 4. Were any incentives offered (e.g., monetary, prizes, or non-monetary incentives such as an offer to provide the survey results)? | 4. Respondents were offered a $25 or $50 Amazon gift card for completing the survey after the first or second invitation, respectively. |
| **5. Time/Date** | 5. In what timeframe were the data collected? | 5. The survey was first launched on April 18, 2021 and was closed on August 27, 2021. |
| **6. Randomization of items or questionnaires** | 6. To prevent biases items can be randomized or alternated. | 6. Respondents were randomly allocated in a 1:1:1:1 ratio to the 4 versions of the survey, which varied the reported patient race in the atherosclerotic cardiovascular disease and prostate cancer case scenarios. |
| **7. Adaptive questioning** | 7. Use adaptive questioning (certain items, or only conditionally displayed based on responses to other items) to reduce number and complexity of the questions. | 7. N/A |
| **8. Number of Items** | 8. What was the number of questionnaire items per page? The number of items is an important factor for the completion rate. | 8. Each page had no more than one question, and multi-part questions had up to 9 parts. |
| **9. Number of screens (pages)** | 9. Over how many pages was the questionnaire distributed? The number of items is an important factor for the completion rate. | 9. The survey spanned 16 pages. |
| **10. Completeness check** | 10. It is technically possible to do consistency or completeness checks before the questionnaire is submitted. Was this done, and if "yes", how (usually JAVAScript)? An alternative is to check for completeness after the questionnaire has been submitted (and | 10. Yes, data completeness was assessed both during and after survey submission. As a dynamic completeness check, each of the first 10 questions (all except those on respondent characteristics) required a response before the respondent could proceed to the next question. |

| | | |
|---|---|---|
| | highlight mandatory items). If this has been done, it should be reported. All items should provide a non-response option such as “not applicable” or “rather not say”, and selection of one response option should be enforced. | After survey submission, the distribution of completed questions was examined and response validity assessed with response timing, longstring analysis, and Mahalanobis distance |
| **11. Review step** | 11. State whether respondents were able to review and change their answers (e.g., through a Back button or a Review step which displays a summary of the respons- es and asks the respondents if they are correct). | 11. Back button and response editing were enabled. |
| **Category: Response rates** | | |
| **1. Unique site visitor** | 1. If you provide view rates or participation rates, you need to define how you determined a unique visitor. There are different techniques available, based on IP addresses or cookies or both. | 1. Duplicate responses were identified using a unique identifier in each survey link. In the case of duplicate responses, the earliest valid response was included for analysis. |
| **2. View rate (Ratio of unique survey visitors/unique site visitors)** | 2. Requires counting unique visitors to the first page of the survey, divided by the number of unique site visitors (not page views). It is not unusual to have view rates of less than 0.1 % if the survey is voluntary. | 2. View rate was 14.7%, defined as the number of unique clickers (409) over the number of email openers (2,776). |
| **3. Participation rate (Ratio of unique visitors who agreed to participate/unique first survey page visitors)** | 3. Count the unique number of people who filled in the first survey page (or agreed to participate, for example by checking a checkbox), divided by visitors who visit the first page of the survey (or the informed consents page, if present). This can also be called “recruitment” rate. | 3. Participation was rate was 96.3%, defined as the number of respondents consenting to the study by clicking to proceed beyond the first page (394) over the number of unique clickers (409). |
| **4. Completion rate (Ratio of users who finished the survey/users who agreed to participate)** | 4. The number of people submitting the last questionnaire page, divided by the number of people who agreed to participate (or submitted the first survey page). This is only relevant if there is a separate “informed consent” page or if the survey goes over several pages. This is a measure for attrition. Note that “completion” | 4. Completion rate was 93.1%, defined at the number of fully completed surveys (367) over the number of consenting respondents (394). |

| | | |
|---|---|---|
| | can involve leaving questionnaire items blank. This is not a measure for how completely questionnaires were filled in. (If you need a measure for this, use the word “completeness rate”.) | |
| **Category: Preventing multiple entries from the same individual** | | |
| **1. Cookies used** | 1. Indicate whether cookies were used to assign a unique user identifier to each client computer. If so, mention the page on which the cookie was set and read, and how long the cookie was valid. Were duplicate entries avoided by preventing users access to the survey twice; or were duplicate database entries having the same user ID eliminated before analysis? In the latter case, which entries were kept for analysis (e.g., the first entry or the most recent)? | 1. Once the survey was opened, a cookie was placed on the user's browser and lasted until the survey was closed or expired. This could be bypassed by clearing cookies or using another browser or incognito window. Unique identifiers were added to each survey link through IQVIA and Qualtrics to check for duplicate responses. |
| **2. IP check** | 2. Indicate whether the IP address of the client computer was used to identify potential duplicate entries from the same user. If so, mention the period of time for which no two entries from the same IP address were allowed (eg, 24 hours). Were duplicate entries avoided by preventing users with the same IP address access to the survey twice; or were duplicate database entries having the same IP address within a given period of time eliminated before analysis? If the latter, which entries were kept for analysis (e.g., the first entry or the most recent)? | 2. IP addresses were not used to identify duplicate entries. |
| **3. Log file analysis** | 3. Indicate whether other techniques to analyze the log file for identification of multiple entries were used. If so, please describe. | 3. N/A |
| **4. Registration** | 4. In “closed” (non-open) surveys, users need to login first and it is easier to prevent duplicate entries from the same user. Describe how this | 4. Duplicate responses were identified using a unique identifier in each survey link. In the case |

| | | |
|---|---|---|
| | was done. For example, was the survey never displayed a second time once the user had filled it in, or was the username stored together with the survey results and later eliminated? If the latter, which entries were kept for analysis (e.g., the first entry or the most recent)? | of duplicate responses, the earliest valid response was included for analysis. |
| **Category: Analysis** | | |
| **1. Handling of incomplete questionnaires** | 1. Were only completed questionnaires analyzed? Were questionnaires which terminated early (where, for example, users did not go through all questionnaire pages) also analyzed? | 1. Fully completed surveys were defined as those in which participants responded to all elements (Q1-Q13). Partially completed responses were defined as those including completion of survey items through and beyond the clinical case scenarios (Q1-Q6), but not completed in full. |
| **2. Questionnaires submitted with an atypical timestamp** | 2. Some investigators may measure the time people needed to fill in a questionnaire and exclude questionnaires that were submitted too soon. Specify the timeframe that was used as a cut-off point, and describe how this point was determined. | 2. Response time <4 minutes was considered as an indicator of careless response, corresponding to the top 5th percentile of response times across all participants. Sensitivity analyses excluding these and other potentially careless responses did not change the data interpretation. |
| **3. Statistical correction** | 3. Indicate whether any methods such as weighting of items or propensity scores have been used to adjust for the non-representative sample; if so, please describe the methods. | 3. N/A |
